# Improving 3D-CINE tTV-regularized whole-heart MRI reconstruction

**DOI:** 10.1101/2024.02.22.24302997

**Authors:** Bastien Milani, Christopher Roy, Jean-Baptiste Ledoux, David C. Rotzinger, Ambra Masi, Renaud Troxler, Salim Si-mohamed, Jerome Yerly, Ludovica Romanin, Tobias Rutz, Estelle Tenisch, Milan Prsa, Juerg Schwitter, Matthias Stuber

## Abstract

**Purpose:** To improve the image quality of 3D radial free-running MRI data of the heart through a deliberate and stepwise extension of the XD-GRASP reconstruction.

**Methods:** Ferumoxytol-enhanced cardiac free-running 3D-radial data were reconstructed using an XD-GRASP reconstruction improved by 4 new developments: motion-compensated temporal-Total-Variation (MC-tTV) regularization for 3D images, a new coil-sensitivity, a new k-space density compensation and a revisited conjugate-gradient-descent (with exact line search) for solving the least-square sub-problem of ADMM. The resulting images were compared quantitatively and qualitatively to reconstructions lacking some of the newly implemented measures. Also, the measurement of ejection-fraction by a threshold-based method on the new reconstruction was compared to a reference standard.

**Results:** The new reconstruction significantly increased the sharpness of the right coronary artery (4% to 6%, p < 0.05) and the left anterior descending coronary artery (4% to 5% p < 0.05). It also increased blood-myocardium interface sharpness (between 20% and 25%, p < 0.05) and decreases spatial-Total-Variation in the blood-pool (13%, p < 0.05). The qualitative evaluation suggests better anatomical depiction of small structures using the new reconstruction. As compared to a reference standard method, ejection fraction could also be correctly evaluated.

**Conclusion:** Compressed sensing image reconstruction for 3D-radial free-running cardiac acquisition was successfully improved by including MC-tTV regularization, a new density compensation, a new coil-sensitivity and a revisited conjugate-gradient-descent with exact line search. Quantitative and qualitative quality metrics demonstrated significant improvement in image quality when using the new reconstruction, while extracted dynamic information compared favorably with the gold standard.

## Introduction

Free-running MRI techniques for three-dimensional (3D) time-resolved whole-heart magnetic resonance imaging (MRI) is an active area of research. Using such techniques and equipped with total self-navigation (1), time efficiency is high, and the paradigm changes from prospective scan planning and scan parameter adjustment to fully flexible and retrospective analysis of the acquired data. Operator dependency may be reduced, 3D CINE images with high temporal and isotropic spatial resolution can be obtained, and no breath-holding neither placement of ECG electrodes are required. Such a free-breathing approach is potentially advantageous for all patients, in particular very sick and dyspneic patients, and avoids in general potential disturbances of hemodynamics that can occur during breath-holding if the patient exerts mild Valsalva-maneuver (2). However, acquisition time constraints combined with binning of cardiac and respiratory phases result in a high degree of under-sampling in Fourier space for each 3D CINE image to reconstruct. As a result, the achievable resolution and scan time is inherently tied to advances in image reconstruction methods. In fact, compressed sensing techniques (CS) and its related precursors (3–32) have already played an important role in Cardiac CINE MRI reconstruction as demonstrated by the existence of an extensive literature. Taking advantage of the sparsity in some domain of the images to reconstruct, high degrees of undersampling beyond the Nyquist sampling limit can be accommodated using CS, which offers new opportunities for improved spatial resolution, temporal resolution, volumetric coverage, and ease of use. These techniques have been successfully applied to 2D-CINE cardiac reconstruction (6–12) as well as to 3D-CINE Cartesian (3–5) and radial reconstructions (15).

Among some of these approaches to cardiac CINE reconstructions, a particularly fruitful strategy is to assume that the difference between temporally adjacent frames should be sparse (which means a high degree of redundancy between temporally adjacent frames) and this has led to the concept of temporal-total-variation (tTV) regularization in MRI reconstruction. CS reconstructions regularized with tTV, in particular the XD-GRASP reconstruction (33), have shown promising results in the domain of cardiac CINE imaging (32). However, an important user-defined trade-off exists where a large regularization weight can introduce a loss of sharpness, a loss of structural information and a compression of motion, while a small regularization weight over-emphasizes the data-fidelity term and may not sufficiently account for under-sampling artifacts.

Strategies against the adverse effects of motion in compressed sensing reconstructions for cardiac CINE MRI has been explored using different methods (13–30). In MASTER, a technique for reconstructing 2D-CINE cardiac MRI (13), deformation fields are used in order to increase sparsity in the difference between adjacent CINE frames providing inter-bin compensation of cardiac motion. The authors of MASTER define the motion-compensated residuals as the motion-compensated difference between temporally adjacent frames, the sum of which will be called “motion-compensated temporal-Total-Variation” (MC-tTV) in the present text. However, MASTER has been implemented for 2D cartesian data only and MC-tTV has never been incorporated to an XD-GRASP reconstruction for 3D-radial data.

The XD-GRASP reconstruction suffers therefore of not being informed by deformation fields since it is regularized by tTV and not MC-tTV. But it is not the only drawback of that reconstruction: the algorithm used to solve the reconstruction problem is a non-linear conjugate gradient descent (CGD) with inexact line search (34), which is not suited for non-differentiable objective functions (the 1-norm is not differentiable). To our knowledge, this drawback has never been completely solved for cardiac MRI reconstructions. More advanced algorithm, such as alternating direction method of multipliers (ADMM), have been proposed for CS reconstructions. But ADMM contains a least-square sub-problem that needs to be solved with CGD, and while CGD has been published originally with exact line-search (35), the MRI literature only still reports inexact line-search (for example (36), (37)) or a detailed account of the line search is not provided (for example (14)). It may therefore be that existing XD-GRASP implementation are not optimal in term of line-search.

Beside the two above mentioned drawbacks of XD-GRASP, the existing implementations of density compensation for 3D-radial data are possibly suboptimal since uniformity of the data lines over the sphere surface has always been assumed, as far as we know, although the binning operation leads to a significantly non-uniform covering of the sphere surface in k-space. Finally, the coil-sensitivity estimation used for 3D-radial cardiac imaging has produced very non-uniform image intensities has it can be seen on figures in the literature (1,13,33).

The purpose of the present article is to address the four mentioned drawbacks of XD-GRASP for 3D-radial cardiac MRI (1,38,39) by four original combined developments. The first is the replacement of tTV in XD-GRASP by MC-tTV. Inspired by MASTER that was demonstrated for cartesian 2D-CINE imaging, we integrated inter-bin compensation of cardiac motion into an XD-GRASP reconstruction for free-running 3D radial data providing a technique for 4D whole-heart imaging. The second development is an implementation of ADDM for XD-GRASP with a revisited implementation of CGD using exact line search. In particular, a mathematical formalism has been developed for that purpose. The third development is a new implementation of the density compensation for 3D-radial trajectories, which makes use of the Voronoi algorithm for taking the non-uniform distribution of data line in k-space into account. The fourth development is a new coil-sensitivity estimation for recovering a spatially homogeneous image intensity in 3D cardiac imaging.

We tested the hypothesis that our four proposed measures significantly improve the quality of images reconstructed from fully self-gated free-running 3D-radial data acquired on a cohort of patients with congenital heart disease after ferumoxytol injection.

## Methods

To address the shortcomings related to the tTV regularization, the CGD algorithm with inexact line-search, the density compensation and the coil-sensitivity estimation for XD-GRASP reconstructions of 3D-radial data, the four following methods have been developed.

i. MC-tTV regularization for 4D cardiac imaging (3D+time) was implemented in an XD-GRASP reconstruction. A mathematical formalism was developed for that purpose and is presented in the supplementary material. Briefly, we call ***x***^(1)^, …, ***x***^(*nFr*)^ the list of 3D frames to be reconstructed, where *nFr* is the total number of frames and where each ***x***^(*i*)^ is written as a column vector by column major order. We call ***DF***^(*i*)^ the deformation-field that deforms frame ***x***^(*i*)^ to match frame ***x***^(*i*−1)^ and which was estimated with a third-part software (see sections below). We implemented a class to store sparse-matrix ***T***^(*i*)^ (and its transpose matrix, for the need of the optimization algorithm) which encodes ***DF***^(*i*)^ and which realizes the corresponding deformation as ***T***^(*i*)^***x***^(*i*)^ ≈ ***x***^(*i*−1)^. A C++ function was also implemented in order to realize the sparse-matrix multiplication with openMP parallelization.
ii. The ADMM algorithm was implemented in order to solve the optimization problem of XD-GRASP with MC-tTV regularization. The least-square sub-problem of ADMM was solved with conjugate-gradient-descent (CGD) using non-standard Euclidean products in order to take care of the density compensation (and other parameters) and to be able to perform exact line-search as in the original publication of CGD (35). We refer to reader to the supplementary material and to an online available document (40) for more details.
iii. A new density compensation for radial trajectories was implemented in order to take care of the non-uniform distribution of data lines over the sphere in k-space and is described in details in the supplementary material. Briefly, the end point of each line (lying on a sphere) was mapped to a plane in a similar (but different) way like stereographic projection. The 2D Voronoi algorithm was then performed for these points (on a plane) and each obtained volume element was mapped back to its corresponding point on the sphere and corrected according to the polar angle.
iv. A new coil-sensitivity estimation was implemented and is described in detail in the supplementary materiel. Briefly, the method is a variant of the one presented in (41). The main difference is that we solve a Laplace boundary value problem to estimate the coil-sensitivity in areas lacking signal, such as the air in the lungs and around the patient body.

To evaluate the individual and combined, cumulative benefit of these four methods (i-iv), different reconstructions were performed on data acquired with a fully self-gated free-running 3D-radial sequence in a cohort of patients with congenital heart disease after ferumoxytol injection. Since combining the presence or absence of these four developments would result in 16 different reconstructions, and thus 120 systematic comparisons, we restricted our study to four coil-sensitivities and three reconstructions. The three reconstruction are the following:

‐ Our new reconstruction consisting of XD-GRASP with MC-tTV (i) and all three other developments (ii-iv). Since it is a 3D version of MASTER by nature, we called it “**MASTER_3D+**” (“MASTER” because it is regularized with MC-tTV (i) and “+” because it contains all other developments (ii-iv)). As a 3D version of MASTER, it is a 4D reconstruction (3D + time) and uses only a quarter of the free-running data (see section below).
‐ Our XD-GRASP reconstruction without MC-tTV but with development (ii-iv) was called “**XD-GRASP_4D+**” (“+” because it contains developments (ii-iv)). It is a 4D reconstruction (3D + time) which uses only a quarter of the free-running data.
‐ As a reference standard reconstruction for comparison, we used the 5D reconstruction developed in Lausanne which already served for several publications. We refer the reader to (1) for a complete definition. It is a 5D reconstruction (3D + cardiac time + respiratory time) and uses the totality of the free-running data. We called it “**XD-GRASP_5D_LAUS**” in the present article.

We organized the comparisons in two studies and we performed an analysis of the convergence of MASTER_3D+ as a third study. We define our three studies as follows:

‐ Study 1: Comparison of MASTER_3D+ versus XD-GRASP_5D_LAUS and versus XD-GRASP_4D+ by the mean of quantitative and qualitative end points. It was also verified in this study if the ejection-fraction (EF) measured with MASTER_3D+ by a threshold based method was in accordance with a gold-standard measurement.
‐ Study 2 **(supplementary material)**: Comparison of our new coil-sensitivity estimation to 3 other coil-sensitivity estimation using quantitative metrics, including the difference of measured EF as compared to a reference standard.
‐ Study 3 **(supplementary material)**: Convergence analysis of MASTER_3D+.

### Data acquisition

The local ethics committee approved the study, and written informed consent was obtained from all individuals or their legal representatives. The acquisition was performed in 12 patients (additional details provided by Table 1) with congenital heart disease on a whole-body 1.5T clinical MR scanner (MAGNETOM Sola, software version VA20A, Siemens Healthineers, Erlangen, Germany). All data were acquired after injection of a 2-5mg/kg dose of ferumoxytol that served to increase the contrast between blood and myocardium (38). A free-running gradient-echo sequence with 3D-radial phyllotaxis sampling trajectory (42) was acquired after conventional 2D-CINE imaging. There acquisition was therefore not gated. Table 2 presents all relevant sequence parameters.

**Table 1:** Table of participants. **The entries of this table have been masked in order to discard identifying information as required for the pre-print version of the article**.

| Patient number | Age (years) | Gender | Diagnosis | Heart Rate (bpm) | Weight (kg) | Height (cm) | Body Surface Area (m<sup>2</sup>) |
| --- | --- | --- | --- | --- | --- | --- | --- |
| 1 |  |  |  |  |  |  |  |
| 2 |  |  |  |  |  |  |  |
| 3 |  |  |  |  |  |  |  |
| 4 |  |  |  |  |  |  |  |
| 5 |  |  |  |  |  |  |  |
| 6 |  |  |  |  |  |  |  |
| 7 |  |  |  |  |  |  |  |
| 8 |  |  |  |  |  |  |  |
| 9 |  |  |  |  |  |  |  |
| 10 |  |  |  |  |  |  |  |
| 11 |  |  |  |  |  |  |  |
| 12 |  |  |  |  |  |  |  |
| Average | 21.3 | 100% male | - | 60.3 | 67.4 | 175 | 1.8 |

**Table 2:** Acquisition parameters of the MRI sequences.

|  | **Free-running Sequence** | **2D CINE Sequence** |
| --- | --- | --- |
| Sequence Type | GRE | BSSFP |
| Dimensionality | 3D | 2D |
| TR | 2.84 ms | 3.06 ms |
| TE | 1.64 ms | 1.25 ms |
| Matrix size | 192 frequency encoding | 208 frequency-enc., 166 phase-enc. |
| Flip Angle | 15 deg | 80 deg |
| Field of View | 220x220x220 mm<sup>3</sup> isotropic | 300 x 240 mm<sup>2</sup> |
| Excitation Type | Slab-selective | Slice-selective |
| Pixel Bandwidth | 1002 Hz/Pix | 925 Hz/Pix |
| Number of Lines per Segment | 22 | 15 |
| Number of Interleaves | 5749 | 11 |
| Spatial Resolution | 1.15 x 1.15 x 1.15 mm<sup>3</sup> | 1.4 x 1.4 x 5.0 mm<sup>2</sup> |
| Scan Time | 6 min | 6 min |

### Image Reconstruction

We refer the reader to the literature (1) for the definition of XD-GRASP_5D_LAUS. It is a 5D reconstruction that makes use of the totality of the free running data. The reconstructions XD-GRASP_4D+ and MASTER_3D+ were performed as follows.

The coil-sensitivities of the surface coil-array elements (18 channels) were estimated following our new method (iv), by the use of low-resolution prescan images acquired with both the body coil of the scanner and surface receiver, as described in the supplementary material. A matrix size of 96x96x96 was used for that estimation. *nCh* will refer to the number of channels (or coils).

The superior-inferior-projections (SI-projections) acquired as the first line of each interleaf (i.e. each shot) were used to extract cardiac and respiratory self-gating signals as described previously (1). These self-gating signals were then used to sort the image data (and corresponding trajectory points) into cardiac and respiratory motion-resolved 5D data sets or bins. Respiratory motion was suppressed by selecting only end-expiratory data resulting in cardiac motion-resolved data sets. Therefore, all reconstructions were performed on end-expiratory cardiac motion-resolved 4D data that account for 25% of the acquired data. Each cardiac bin was selected in such a way that it covered a duration of 50 ms.

K-space data of the *i*th bin in the cardiac cycle will be written as ***y***^(*i*)^, which corresponds to image frame ***x***^(*i*)^. The vertical concatenation ***y*** = [***y***^(1)^; … ; ***y***^(*nFr*)^] represents the complex valued data. We note *Y* the vector space that contains *y* and *Y*^(*i*)^ the vector space containing ***y***^(*i*)^.

A gridded reconstruction for each frame was achieved by a non-uniform Fourier-inverse on each data bin followed by a coil combination (Moore-Penrose pseudo-inverse). To ensure that similar regularization weights can be used across different subjects, the average magnitude value was estimated in the ventricular blood pool of the first frame by manually drawing an ROI and all the data bins were then normalized to this value. This way, raw data were rescaled to obtain the ensuing reconstructions with a blood-pool value close to 1. The present gridded reconstruction was rescaled accordingly and was used as the initial image for the CS-reconstructions.

The optimization problem for both **XD-GRASP_4D+** and **MASTER_3D+** is the generalized-LASSO problem

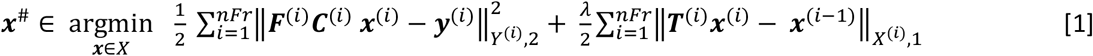

where 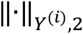 is the L2-norm on space *Y*^(*i*)^ that contains the k-space data ***y***^(*i*)^ of frame number *i*, 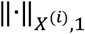 is the L1-norm on the space *X*^(*i*)^ that contains frame ***x***^(*i*)^, ***C***^(*i*)^ is the coil-sensitivity map of frame number *i*, ***F***^(*i*)^ is the non-uniform Fourier transform from 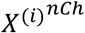 to *Y*^(*i*)^ and *λ* is a real regularization parameter. For XD-GRASP_4D+, each matrix ***T***^(*i*)^ is simply the identity while for MASTER_3D+ each ***T***^(*i*)^ is a linear map that performs a non-rigid deformation of ***x***^(*i*)^ in order to align it with ***x***^(*i*−1)^. In the former sum, we use the convention that index *i* = 0 is replaced by *i* = *nFr* in a circular manner.

Of note, the different ***C***^(*i*)’^s are all copies of the same matrix in the current implementation. The coil sensitivities are therefore time-independent and identical for all frames. Nevertheless, discriminating them by different symbols allows for a more general mathematical description that may be of interest for future reconstructions.

The above optimization problem [1] is an instance of the generalized-LASSO problem. It was solved with the ADMM algorithm (Alternating Direction Method of Multipliers) (43). Our new density compensation (iii) served as diagonal elements of the matrix defining the inner product on each *Y*^(*i*)^ while the matrix defining the inner product on each *X*^(*i*)^ was set as the voxel size times the identity. Our new density compensation (iii) is thus part of the definition of each 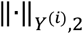 and is also part of the adjoint operator involved in the conjugate-gradient-descent (CGD) for solving the least-square sub-problem of ADMM (see supplementary material). The use of these non-standart inner products allows to perform exact line-search in the CGD, which is a key element to our new developpment (ii).

The reconstruction **XD-GRASP_4D+** was performed by solving [1] with *λ* = 0.1 and setting 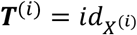 (the identity on each frame-space *X*^(*i*)^), resulting in a conventional tTV-regularization. The deformation field ***DF***^(*i*)^ (see supplementary method) between each frame ***x***^(*i*)^ and its previous temporal neighbor ***x***^(*i*−1)^ (obtained from XD-GRASP_4D+) was then estimated using the NiftyReg registration software (44,45), considering that ***x***^(*nFr*)^ precedes ***x***^(1)^ in circular manner. Each ***DF***^(*i*)^ was transformed into a sparse deformation matrix ***T***^(*i*)^ being the gridding matrix on the new position given by ***DF***^(*i*)^. This means that, if the registration process and XD-GRASP_4D+ were perfect, it would hold that ***T***^(*i*)^***x***^(*i*)^ = ***x***^(*i*−1)^. **MASTER_3D+** was finally performed as a second iterative reconstruction by choosing *λ* = 0.3 and setting ***T***^(*i*)^ to be the *i*th deformation matrix that deforms frame ***x***^(*i*)^ in order to match frame ***x***^(*i*−1)^. The use of deformation fields in MASTER_3D+ and the implementation of the code for that purpose is our new development (i).

The ADMM parameter *ϱ* ((43) page 14) was chosen to be 10*λ*. The regularization parameters (*λ* = 0.1, *λ* = 0.3 and *ρ* = 10 *λ*) were chosen experimentally by repeating the reconstruction on one data set and by visually assessing image quality. The reconstruction matrix size was 384 x 384 x 384 and the reconstruction field of view was 440 mm isotropic leading to an isotropic voxel size of 1.15 mm.

All reconstructions were run on a cluster computer equipped with a job-scheduling system (SLURM). No heavy parallelization was used (i.e., no work distribution between nodes). Each reconstruction was performed on one of the available nodes (2x AMD EPYC 7742 @ 2.25GHz with 64 cores = 128 CPUs, 2TB RAM, 2 nVidia RTX 3090 24GB GPU) which may have been a different one for different reconstructions. Depending on resource availability, 32 to 48 cores were selected. A maximum of 60 ADMM iterations containing each 3 conjugate-gradient-descent (CGD) iterations was used. The stopping criteria for the reconstruction was that the maximum number of iterations was reached, or that the maximum available computation time (48 hours) was reached. The reconstruction time was recorded for every reconstruction.

The reconstruction was implemented in MATLAB (MathWorks, Natick, Massachusetts, USA) except for the gridding operations of the non-uniform Fourier transform ***F***^(*i*)^ and of the image deformations ***T***^(***i***)^, which were coded in C++ and compiled as MATLAB-executable-files (MEX-files) and for which CPU parallelization was implemented by means of OpenMP.

### Study 1: Comparison of MASTER_3D+ versus XD-GRASP_4D+ and XD-GRASP_5D_LAUS

The purpose of study 1 is to determine whether the four additional developments present in MASTER_3D+ leads to improved image quality relative to XD-GRASP_4D+ and XD-GRASP_5D_LAUS. Another purpose is to determine whether MASTER_3D+ leads to accurate assessment of ventricular function relative to reference standards. We performed the following analyses.

#### Quantitative image comparison metrics

We define in this subsection quantitative metrics (or endpoints) for images comparison. For measurements related to the heart, those quality metrics were extracted on end-diastolic images as determined by the cardiologist.

Left anterior descending artery (LAD) and right coronary artery (RCA) vessel sharpness and traceable vessel length were measured with Soap-bubble (46). Note that coronary vessel sharpness is used as a surrogate end-point for image quality and quantitatively informs about the quality of motion suppression for a given approach.

The interface sharpness between blood pool and myocardium (BMIS for “blood-muscle interface sharpness”) was measured with a Matlab tool (47). Given two reconstruction *A* and *B*, the relative sharpness gain (in %, positive or negative) was calculated as

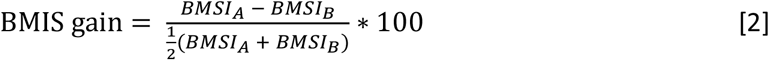

To quantify a reduction (positive or negative) in noise-like errors (including a combination of noise, reconstruction error and artifacts) between two reconstruction *A* and *B*, a 2-dimensional region of interest (ROI) in a homogeneous region of the blood-pool (free of flow artifacts) was selected manually on a coronal plane (plane number *k*). Symbol *x*_*ijk*_ will stand for the voxel value of row *i*, column *j*, and plane *k* of a 3D image. The symbol “*x*_*ijk*_ ∈ *ROI*” will mean that the voxel of row *i*, column *j*, and plane *k* is located inside the given 2D-ROI. The spatial (not temporal) total-variation (sTV) inside that 2D-ROI will be written *sTV*^*ROI*^ and was calculated as

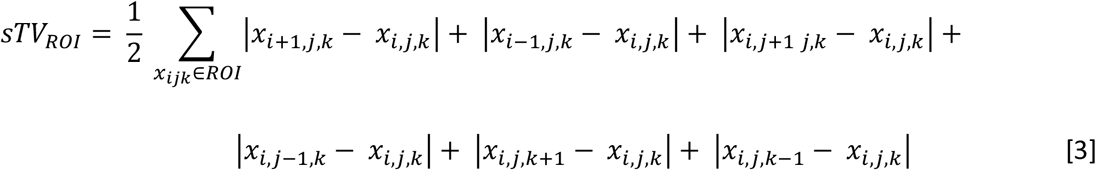

The sTV reduction (in %) between reconstruction *A* and *B* was calculated as

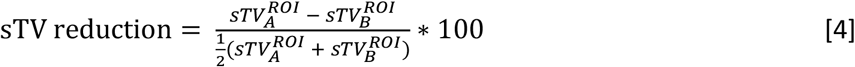

Similarly, the ratio between the average blood signal and its standard deviation in the same ROI was computed and written as the symbol “[*µ/σ*]^*ROI*^ “. Because CS images do not exhibit a regular (Gaussian or Riccian) noise, we named this ratio “*µ/σ*-ratio” and not “signal-to-noise”. The relative *µ/σ* gain between any two reconstruction *A* and *B* (in %) was evaluated as

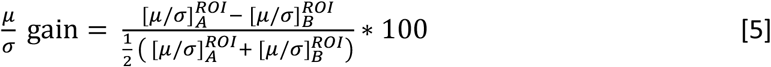

The significance of an average (over patients) being different from 0, or an average difference being different from 0, was assessed by a two-tailed t-test for any quantitative end-point defined above.

We define here the EF-bias of a given reconstruction as the average difference (over all patients) between the EF measured with that reconstruction and the EF measured with the reference standard method using 2D-CINE imaging. The EF-bias (given as mean±SD) expresses thus the accuracy of EF of the various reconstructions. On the reference 2D-CINE images, EF was measured using commercial software by a reader blinded to the 3D-CINE results and with 5 years of CMR analysis experience (AM). On 3D-CINE reconstructions, EF was measured blinded as follows.

End-systolic and end-diastolic cine frames were identified by the cardiologist. Then, the three following masks were determined on both end-systolic and end-diastolic cine frames:

‐ a manually drawn mask with boundaries lying inside the LV myocardium (inside the muscle),
‐ one 3D half-space (the set of points lying on one side of a plane) separating the LV from the left atrium,
‐ another 3D half-space separating the LV from the aortic valve.

The intersection of these three masks resulted then in a “shell-mask” that contains the LV (mainly blood and some muscle) and excludes the rest of the image. To segment the blood inside the shell-mask, a threshold-mask was computed for the blood-pool signal. The intersection of this “threshold-mask” and the “shell-mask” resulted in the segmentation of the blood in the LV. The stroke volume was subsequently computed by subtracting the end-systolic from the end-diastolic blood volume. EF (in %) was calculated as the ratio between stroke volume and the end-diastolic volume and multiplied by 100%. The three masks defined by the three bullet points above were determined from the MASTER_3D+ reconstruction showing the better BMIS and were reused for all other reconstructions to have a fair comparison with respect to segmentation, the goal of the present EF comparison being to compare the performance of signal-value-thresholding for EF measurement. The definition of these three masks was blinded to the EF measurement with the reference 2D-CINE method.

Blant-Altman-plot and correlation-plots between EF measured from MASTER_3D+ versus EF measured from the reference standard method were also performed.

#### Qualitative image comparison methods

A blinded qualitative comparison was performed by two experienced (> 10 years and 5 years) radiologist (DR and RT) between the reconstruction MASTER_3D+ versus XD-GRASP_5D_LAUS and also between the reconstruction MASTER_3D+ versus XD-GRASP_4D+ using the following 9 comparison criteria: visual sharpness of left main artery (LM), of the left anterior descending artery (LAD), of the circumflex artery (LCx), of the right coronary artery (RCA) and of the aortic arch branches interface, conspicuity of aortic valve leaflets, perceived noise, overall diagnostic confidence, and Likert scale (48). For each criterion, the following analysis was performed: for each patient, the radiologist recorded which of the two reconstructed images (MASTER_3D+ and one of the other) was better according to the given criteria. The number of patients for which one reconstruction was preferred over the other was reported. The result’s significance was assessed with a two-sided binomial test. The analysis was repeated independently by two radiologists.

Another qualitative analysis was conducted on so-called M-mode images. For each reconstruction was an M-mode image constructed as follows: From each frame of the reconstruction, one line of voxels (direction left-right w.r.t patient body) was chosen. Each line (one per frame) was plotted as a column and concatenated from left to right (i.e. in the time dimension) to obtain a 2D image called M-mode. For each patient, the three M-modes (one for each reconstruction) were displayed next to each other in a random order and the conspicuity of graphical features was compared visually by the author himself.

### Study 2: Comparison of different coil-sensitivity estimations

Study 2 is defined in the supplementary material. It is a study about coil-sensitivities. We compare our new coil-sensitivity estimation to three other methods, in particular to the coil-sensitivity estimation used in XD-GRASP_5D_LAUS.

### Study 3: Convergence analysis

Study 3 is a convergence analysis of our new reconstruction MASTER_3D+. It is also defined in the supplementary material. In particular, we test our reconstruction on simulated data using the XCAT phantom (49–51).

### Additional reconstructions

We also propose in the supplementary material some additional reconstructions in order to answer some legitimate critics about our methodology.

## Results

### Study 1: Comparison of MASTER_3D+ versus XD-GRASP_4D+ and XD-GRASP_5D_LAUS

Table 3 summarizes all quantitative results of study 1 (excepted EF-bias). The RCA of patient 1 and the LM of patient 2 could not successfully be identified, consistent with a known coronary anomaly in those patients. These two arteries were therefore excluded from the analysis. This table shows a significant improvement of vessel sharpness of RCA and LAD in MASTER_3D+ as compared to the other reconstructions, as well as significant BMIS-gain and *µ/σ*-gain.

**Table 3:** Quantitative comparison of **MASTER_3D+** vs **XD-GRASP_4D+** and of **XD-GRASP_5D_LAUS**. NS means “non-significant”.

|  | **MASTER_3D+ vs**<br>**XD-GRASP_4D+** | **MASTER_3D+ vs**<br>**XD-GRASP_5D_LAUS** |
| --- | --- | --- |
| Improvement in vessel sharpness<br>for RCA (Soapbubble) | **6.0 ± 5.2 % (p < 0.05)** | **4.2 ± 3.9 % (p < 0.01)** |
| Improvement in vessel sharpness<br>for LAD (Soapbubble) | **3.8 ± 3.8 % (p < 0.05)** | **4.8 ± 7.1 % (p < 0.05)** |
| Improvement in traceable vessel<br>length for RCA (Soapbubble) | p=NS | p=NS |
| Improvement in traceable vessel<br>length for LAD (Soapbubble) | p=NS | **1.5cm ± 1.9cm (p < 0.05)** |
| BMIS gain | **24.5 ± 23.8 % (p < 0.05)** | **21.3 ± 13.9 % (p < 0.01)** |
| sTV reduction | **12.6 % ± 7.9 % (p < 0.01)** | p=NS |
| $\frac{\mu}{\sigma}$ gain | **8.8 % ± 4.8 % (p < 0.05)** | **27.8 % ± 29.2 % (p < 0.01)** |

Figure 1 displays a reformatted image of the right coronary artery from patient 3 at two different time points where a lot of motion occurs (ventricular contraction and dilatation) and demonstrate that the use of deformation fields leads to better visibility of small anatomical structures.

**Figure 1:**
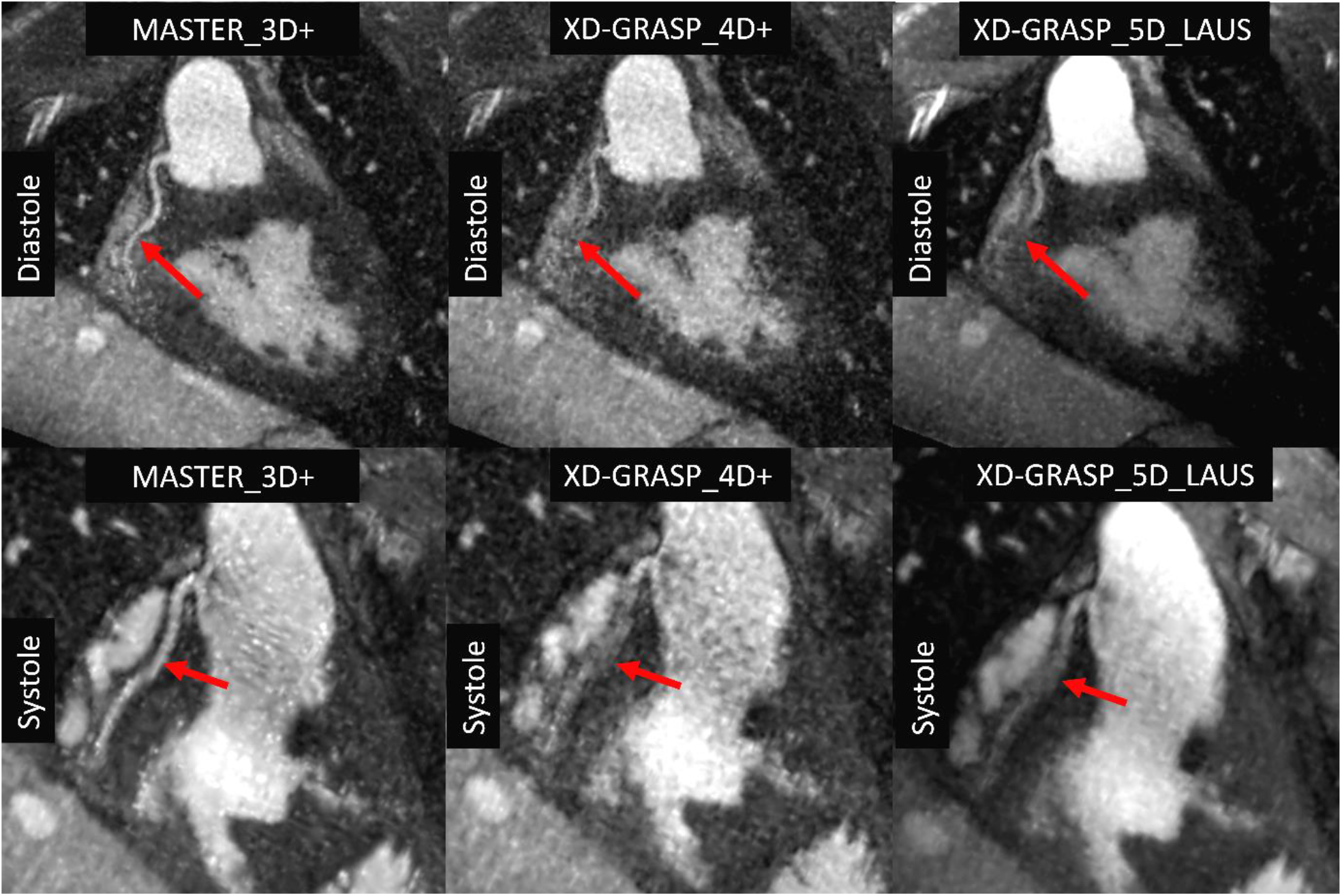
These images were reformatted with SoapBubble and red arrows depict the right coronary artery. The top row corresponds to diastole while the bottom row images originates from systole. MASTER_3D+ (left column) exhibits a better visibility of the right coronary artery as compared to XD-GRASP_4D+ (middle column) and XD-GRASP_5D_LAUS (right column).

Figures 2 shows planes in the three orientations of the reconstructed image of patient 1. Both XD-GRASP_4D+ and XD-GRASP_5D_LAUS show a degradation of moving structures pointed out by red arrows, as compared to MASTER_3D+. One may notice the so-called “salt and pepper” noise on those images (typically in the blood pool).

**Figure 2:**
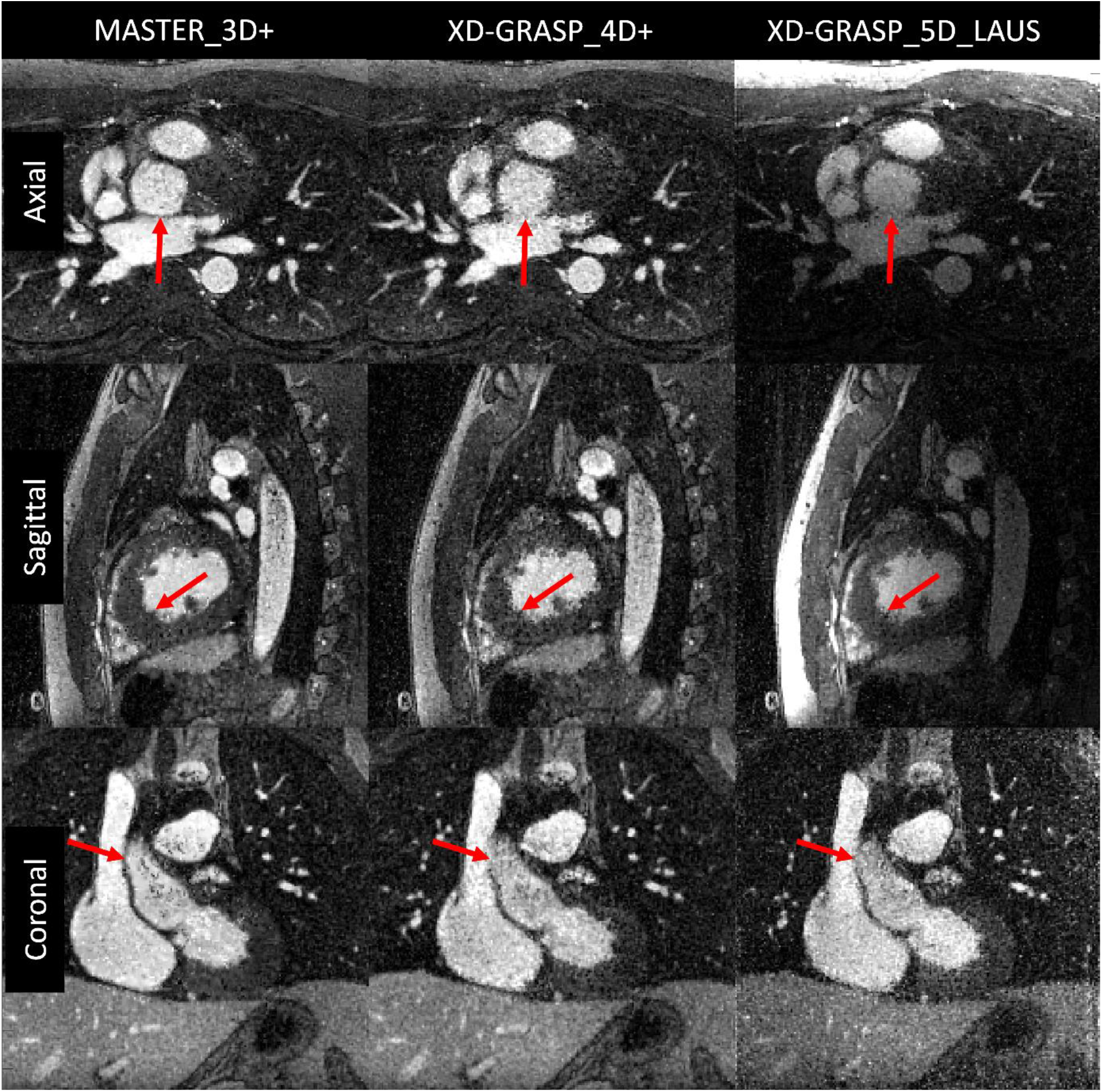
Reconstructed images from data of patient 1. The first raw belongs to diastole while middle and bottom belong to systole. The superiority of MASTER_3D+ over both XD-GRASP_4D+ and XD-GRASP_5D_LAUS can visually be appreciated on details depicted by red arrows (first row, posterior wall of the ascending aorta; second row, left ventricular endocardium and trabeculations; third row, superior vena cava/ascending aorta interface).

Figure 3 shows an example of a moving anatomy before and after deformation, as well as the corresponding deformation field in transverse plane. The registration residuals after deformation show how sparsity improve in motion-compensated residuals.

**Figure 3:**
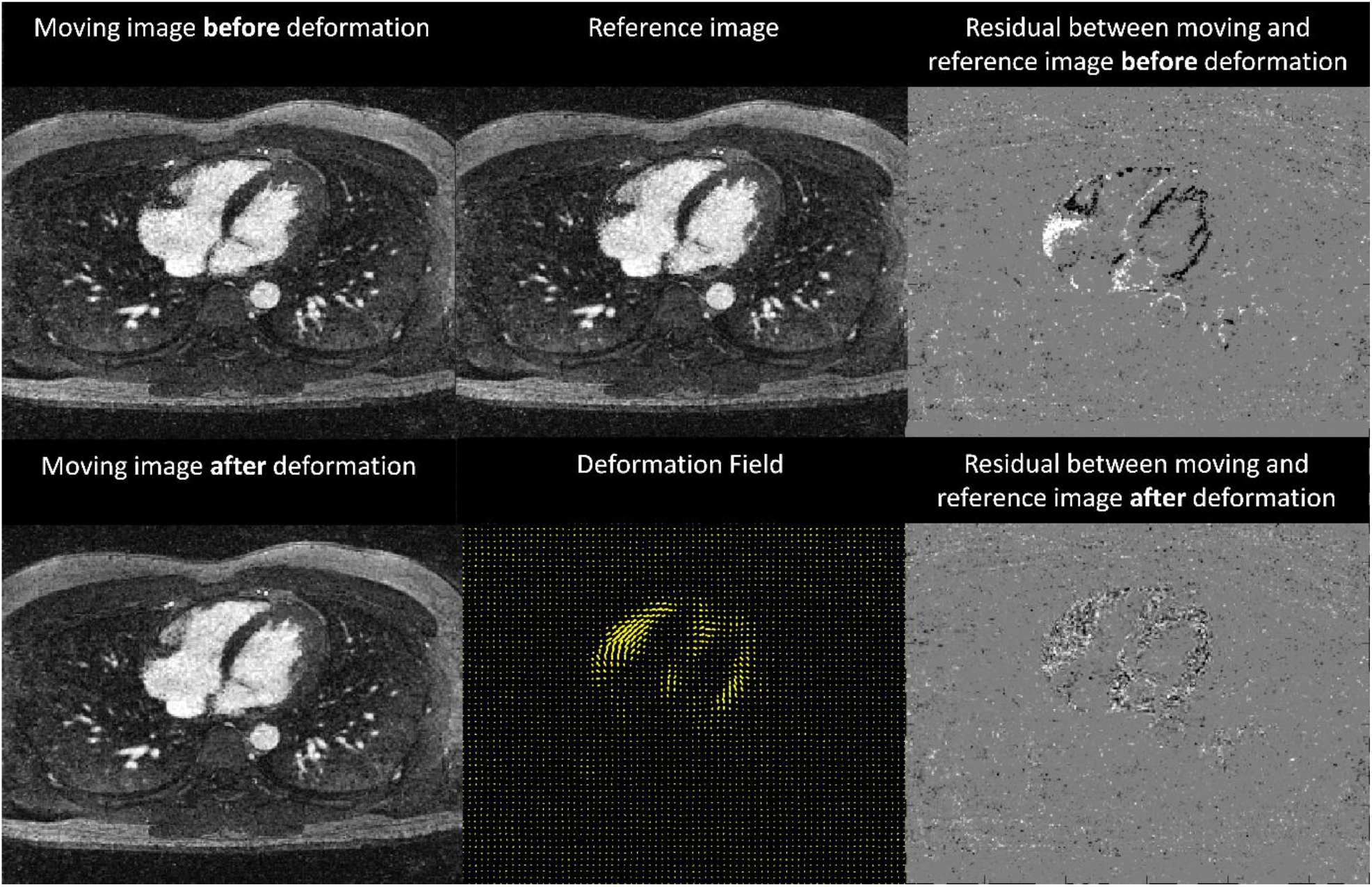
Displayed is an example of moving anatomical image before and after deformation, as well as the corresponding deformation-field in transverse plane. The registration residuals after deformation shows how sparsity improves in the motion-compensated residuals.

Table 4 summarizes the blinded qualitative analysis of the radiologists. Note that the RCA of patient 1 and LM of patient 2 were excluded from the analysis. It shows that none of the evaluated features are better in XD-GRASP_4D+ nor in XD-GRASP_5D_LAUS. Moreover, 5 features were significantly preferred in MASTER_3D+ as compared to XD-GRASP_4D, and 5 other features were significantly preferred in MASTER_3D+ as compared to XD-GRASP_5D_LAUS.

**Table 4:** Blinded qualitative analysis by two radiologists. The left column of top table describes the comparison criteria. The rest of top table displays how many times **MASTER_3D+** was preferred against the compared reconstruction (**XD-GRASP_4D_DF+** or **XD-GRASP_5D_LAUS**) according to the comparison criteria on the left. The bottom table displays the average Likert scale difference between **MASTER_3D+** and **XD-GRASP_4D+** (on the left) resp. **XD-GRASP_5D_LAUS** (on the right).

| Multiple criteria binary comparison performed by two radiologists |  |  |  |  |
| --- | --- | --- | --- | --- |
| Comparison criteria | Number of patients for which **MASTER_3D+** was preferred |  |  |  |
|  | vs **XD-GRASP_4D+** |  | vs **XD-GRASP_5D_LAUS** |  |
|  | radiologist 1 | radiologist 2 | radiologist 1 | radiologist 2 |
| Perceived Noise | **12/12\*\*** | **12/12\*\*** | **10/12\*** | **12/12\*\*** |
| Sharpness of aortic arch branches interface | 8/12 | 12/12** | **10/12\*** | **12/12\*\*** |
| Sharpness of aortic valve leaflets | 9/12 | 10/12* | 8/12 | 7/12 |
| Sharpness of LM (left main artery) | **11/11\*\*** | **10/11\*** | **10/11\*** | **10/11\*** |
| Sharpness of LAD (anterior descending artery) | **12/12\*\*** | **12/12\*\*** | 7/12 | 8/12 |
| Sharpness of LCx (circumflex artery) | **11/12\*\*** | **12/12\*\*** | **10/12\*** | **12/12\*\*** |
| Sharpness of RCA (right coronary artery) | 9/11 | 11/11** | 7/11 | 9/11 |
| Overall diagnostic confidence | **12/12\*\*** | **12/12\*\*** | **12/12\*\*** | **12/12\*\*** |
| Likert scale difference between **MASTER_3D+** |  |  |  |  |
| vs **XD-GRASP_4D+** |  | vs **XD-GRASP_5D_LAUS** |  |  |
| radiologist 1 | radiologist 2 | radiologist 1 | radiologist 2 |  |
| **1.1\*\*** | **1.0\*\*** | **1.1\*\*** | **1.3\*\*** |  |

\*p-value < 0.05
\*\*p-value < 0.01

Four patients were excluded from the EF measurements due to single ventricle anatomy, making the segmentation difficult. As shown in table 5, EF-bias relative to gold standard 2D CINE was 0.0% ± 1.9% for MASTER_3D+ and 0.12% ± 1.6% for XD-GRASP_4D+. But these two biases were not statistically different from 0, nor different between each other. The EF-bias was −3.0% ± 1.8% for XD-GRASP_5D_LAUS, which was significantly different from 0 (p < 0.01). Figure 5A displays EF measured with free-running acquisition and MASTER_3D+ versus EF measured conventionally on 2D-CINE images. Figure 5B shows EF measured conventionally on 2D-CINE images by an observer versus another observer, and figure 5C shows the Bland-Altman plot for the comparison of EF between MASTER_3D+ and the conventional 2D_CINE method.

**Table 5:** All results in the present table are presented in the form “average ±standard-deviation”. The asterisk (*) in row “EF-bias” designates an average significantly different from 0. The asterisk (*) in rows “Std-periphery” and “LLIS” designates an average difference (with other column given inside parenthesis) significantly different from 0. The same results are presented graphically in supplementary figure S5.

| **Coil-Sensitivity** | **A** | **B** | **C** | **D** |
| --- | --- | --- | --- | --- |
| EF-bias (%) | 0.0%±1.9% | -2.13%±2.23%* | 1.50%±1.60%* | 1.38%±1.30%* |
| Std-periphery (arb. units) | 0.48±0.06<sup>*(b)</sup> | 0.74±0.11<sup>*(a, c, d)</sup> | 0.44±0.06<sup>*(b)</sup> | 0.48±0.07<sup>*(b)</sup> |
| LLIS (arb. units) | 2.72 ±0.61<sup>*(c, d)</sup> | 2.77±0.41<sup>*(c, d)</sup> | 1.96±0.45<sup>*(a, b)</sup> | 1.93±0.56<sup>*(a, b)</sup> |

The second part of the qualitative analysis, residing in the inspection of M-mode images, revealed that sharper features in MASTER_3D+ could be found for every patient, as compared to the two non-motion informed reconstructions. The opposite never happened. Figure 4 shows M-mode images with an anatomical plane showing where the M-mode image originates from (yellow line). Top and bottom row correspond to two different patients. One can appreciate how the M-mode images from MASTER_3D+ (B and H) expresses sharper lines as for XD-GRASP_4D+ (D and J) and for XD-GRASP_5D_LAUS (F and L). Red arrows point to examples of such differences. The blur or loss of sharpness in M-mode images is a typical symptom of temporal regularization. Subfigure B and H experimentally demonstrates how this drawback can be corrected.

**Figure 4:**
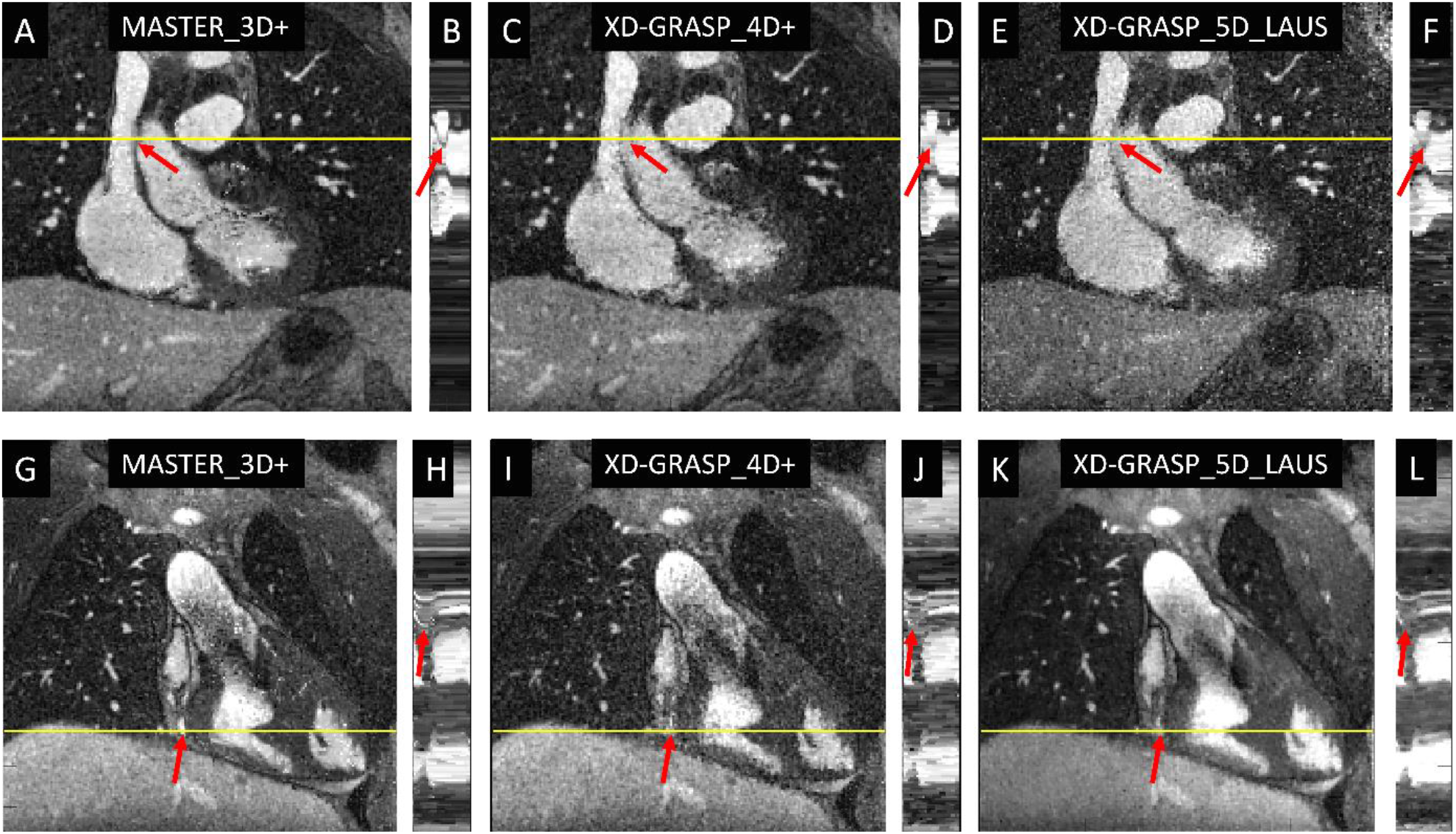
First and second rows display two examples (from two different patients) of M-mode images together with an anatomical plane showing where the M-mode image originates from: the yellow line on the anatomical plane shows the left-right line of voxel chosen to evaluate the M-mode image. Left column (A, B, G, H) corresponds to MASTER_3D+, middle column (C, D, I, J) corresponds to XD-GRASP_4D+, and right column (E, F, K, L) corresponds to XD-GRASP_5D_LAUS. The M-mode images from MASTER_3D+ (B and H) shows sharper features than those from XD-GRASP_4D+ (D, J) and XD-GRASP_5D_LAUS (F, L). Red arrows indicate superior vena cava/ascending aorta interface (upper row), and right coronary artery (lower row).

**Figure 5:**
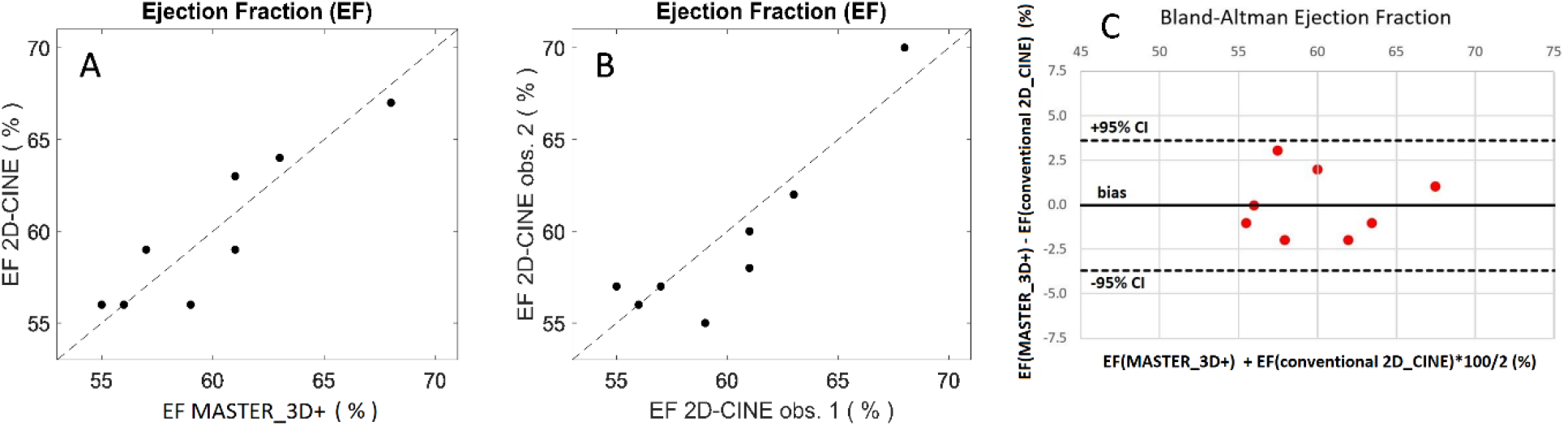
A: Ejection-fraction measured with free-running acquisition and MASTER_3D+ versus Ejection-fraction measured conventionally on 2D-CINE images. B: Ejection-fraction measured conventionally on 2D-CINE images by an observer versus another observer. C: Bland-Altman plot for the comparison of EF between MASTER_3D+ and the conventional 2D_CINE method.

The number of reconstructed cardiac frames was 20.3 ± 4.6. The reconstruction time amounted to 19.4 ± 4.9 hours for XD-GRASP_4D+ and 32.7 ± 9.2 hours for MASTER_3D+ (p<0.05) and the total number of 60 iterations was reached for all patients for those two reconstruction. The reconstruction time was 10.1 ± 2.2 for XD-GRASP_5D_LAUS, which performed 10 iterations for each patient with 4 CGD iterations in each.

### Study 2

The results of study 2 can be found in table 5. We refer the reader to the supplementary material for the complete results of study 2. Briefly, our new coil-sensitivity estimation results in images which are spatially homogeneous and for which EF measurement is not biased as compared to the reference standard method (table 5 and supplementary figures S4 and S5). In contrast, the coil-sensitivity estimation used for XD-GRASP_5D_LAUS leads to spatially non-homogenous images for which the measurement of EF is biased as compared to reference method.

### Study 3 and additional reconstructions

We refer the reader to the supplementary material for the results of study 3. Briefly, the convergence analysis shows that MASTER_3D+ is converging. The results of the additional reconstruction are also presented in the supplementary material.

## Discussion

The quantitative and qualitative comparison between MASTER_3D+ and XD-GRASP_5D_LAUS tested the hypothesis positive that the cumulative effect of motion-compensated temporal-total-variation (MC-tTV) regularization, a revisited conjugate-gradient-descent (CGD) with exact line-search, our new coil-sensitivity estimation and our new density compensation, significantly improves the global quality of images reconstructed from fully self-gated free-running 3D-radial data acquired with ferumoxytol injection. In particular, the visibility of moving structures, signal-to-noise surrogates, and sharpness of coronary arteries were improved. Moreover, the potential problem of cardiac motion being frozen by temporal regularization, and potentially inducing a wrong measurement of ejection fraction, could be rule out. In fact, our EF measurements on MASTER_3D+ were consistent with the gold standard measurement (as shown in figure 5) and suggest that cardiac dynamics can be assessed reliably.

Taken alone, the reduction of sTV in a homogeneous blood pool region, as well as the relative *µ/σ*-gain, do not necessarily suggest improvement in image quality since the same result could have directly been obtained by spatial blurring (filtering) of these images. However, and together with the concomitant sharpness improvements (in RCA, LAD, blood-myocardium interface, M-mode images) without the blurring that typically originates from XD-GRASP_4D+, it may be deduced that the image quality is increasing for MASTER_3D+. Consistent with those quantitative findings, the blinded radiologist reader preferred MASTER_3D+ for most of the evaluated anatomical features. Among the end-points with non-significant p-values, the sharpness of aortic arch branches interface is the least relevant since these vessels are not the main target when performing cardiac MR and likely because they are less subject to motion and may therefore not benefit as much from the motion field reconstructions. Furthermore, aortic valve leaflet conspicuity was not significantly better on MASTER_3D+, which may be linked to the intrinsically high contrast between the leaflets and the blood pool, possibly requiring larger sample sizes to document the effect on small anatomical structures.

Because too many comparisons would have been needed to decouple the effect of each of the four measures individually, we only tested how the image quality decreased when two of them (one at a time) are removed. Namely, we compared the image quality of MASTER_3D+ to the image quality obtained by removing MC-tTV, which results in XD-GRASP_4D+. And in study 2 (supplementary material), we compared the image quality of MASTER_3D+ to that obtained by removing our new coil-sensitivity estimation and replacing it with the one used in XD-GRASP_5D_LAUS (and also two others). The removing of MC-tTV, which consists in XD-GRASP_4D+, results in a significant decrease in image quality as indicated by the quantitative and qualitative analysis, but do not affect the EF measurement. Replacing our new coil-sensitivity by the one used in XD-GRASP_5D_LAUS did not decreased sharpness (see supplementary results) but it changed the overall image homogeneity drastically and created a bias in EF measurement as compared to the gold standard, as shown in supplementary figure S5A.

The effect of the revisited CGD with exact line-search and the new density compensation were not explored individually. We hypothesized in this study that they could be beneficial or, in the worst case, do not damage image quality nor reconstruction time. But for the moment, their individual contribution is confounded in the combination of the four developments (i-iv).

In study 3 (supplementary material), the evolution of the monitor images, the data-fidelity term and the MC-tTV along the 60 iterations, demonstrate experimentally that our reconstruction converges even after a large number of iterations. The decay to zero of the error between reconstructed image and ground-truth for the numerical phantom confirms the convergence of our reconstructions.

Among the four novelties of our work, the most impactful is probably the introduction of motion-compensated residuals between adjacent frames for 3D-radial free-running dynamic MRI, the sum of those residuals being the MC-tTV. Our method is similar to MASTER (13), the main difference being that MASTER was implemented for 2D Cartesian trajectories only, and includes both forward and backward motion-compensated residuals. While we extended this idea to 3D, we implemented only backward residuals in the current implementation to reduce reconstruction time but adding forward residuals can be done straightforwardly. Apart from that difference, we can consider that our method is a 3D version of MASTER applicable to radial acquisitions. The method k-t-FOCUSS with motion correction (21) could be a candidate for comparison to our method. But this was also implemented for 2D Cartesian MRI only. A comparison with k-t-FOCUSS would need to extend it to 3D non-cartesian trajectories. This is however not straight forward because the entire code would have to be re-written to handle large size data and non-cartesian gridding operations. This may be the subject of future studies. The same remark applies for BLOSM (24) and k-t-SLR (52) which exploit block-wise low-rank sparsity, which is another interesting type of regularization.

The reconstruction time of MASTER_3D+ is too high for clinical use, but it was in the intention of the present study to perform the maximum possible number of iterations to explore how fast images would converge. Different strategies could now be pursued to accelerate the reconstruction, including the exploration of a fewer number of iterations, a smaller reconstruction field of view, a multiresolution strategy, and the parallelization of the code among several computer nodes. We also note that the reconstruction time was different among patients since the number of reconstructed cardiac frames depends on individual heart rate, and on the different number of nodes of the HPC that were accessible for the different reconstructions, which may be another confounder.

Our reconstruction with MC-tTV, revisited conjugate CGD with exact line-search, new coil-sensitivity and new density compensation, contributes to the improvement of free-running acquisitions in cardiac MRI, with multiple advantages as compared to the conventional 2D-CINE ECG-gated approach: no ECG and no prospective planning is needed, binning is retrospectively done and can be retrospectively adjusted until a satisfactory result is reached, the full 3D and temporal information becomes available, and finally, no breath-holds are needed. If the reconstruction time can be reduced, the proposed approach would be clinically advantageous for applications that needs a high 3D resolution. In fact, 3D anatomy and function could then be observed for any clinical purpose. As an example, the identification of supracardiac partial anomalous pulmonary vein return, which requires the identification of the connection between the superior vena cava and the right superior pulmonary vein. Another example is the identification of an atrial septal defect, which is often hard to depict properly. A third example is the visualization of the relationship between the coronary artery and the aorta, as well as between the coronary artery and the pulmonary walls when it has an interarterial course.

We note however several limitations in our study. First, ferumoxytol was used as a contrast agent because this was part of the clinical protocol for the acquired data available to test our reconstruction. In principle, our reconstruction should also work with native contrast if the MRI sequence used for acquisition can generate a sufficient contrast (which is not the case for GRE without contrast agent). Yet, this still would have to be tested in practice. Second, our study suffers from an absence of ground truth for deformation-fields and we have no way to estimate them accurately. We can therefore not identify how accurate is the estimation of deformation fields. Even with on simulated data, for which the image ground truth is known, the deformation-fields are difficult to estimate because those images are piece-wise constant, which makes them difficult to register. Third, only a quarter of the data was used by selecting only one respiratory phase. This choice was made to simplify the reconstruction and to build an intermediary (in our opinion necessary) step before using all data for a 5D reconstruction. Fourth, the presented reconstruction technique was tested in patients with regular heart rhythm. How this reconstruction will perform in patients with extrasystoles or other arrhythmias needs further evaluation. Fifth, patients that undergo cardiac MRI often suffer from heart failure which results in irregular breathing patterns (e.g. Cheyne-Stokes breathing). Further testing of the presented reconstruction in these patient populations is warranted. Another limitation is that we don’t have gold standard measurements for coronary imaging (i.e. cartesian gated, fully sampled, fat suppressed, T2 prepared, BSSFP sequence) because that was not part of the clinical protocol. Finally, we note that other temporal regularization strategies (such as temporal Fourier transform or wavelet transform) could have been used in order to reconstruct images prior to deformation-field estimation. These strategies could be explored in a next study.

As a last point of discussion, we note the presence of some spatially isolated bright voxel in MASTER_3D+ (ex. on figure 2). We hypothesize that it is symptomatic of tTV-regularization and may be part of what is sometimes called “salt and paper noise”. Our interpretation is the following. If a voxel is dark like its spatial neighborhood, but that the same voxel location has bright signal on the temporal neighboring frames, it may sometimes happen that temporal regularization enforce a bright value for this voxel in order to mimic its temporal neighbors, without enforcing the same for its spatial neighbors because they have slightly different values and slightly different temporal neighbors. An example of such case can be flow artefacts where some voxels are dark on one frame because of high flow and are bright on the next frame because flow is smaller. Although we claim that flow artefact are candidates for such events, it is only an example and it could potentially happen everywhere on the image where a strong temporal variation of signal exists. In fact, we see these white dotes mostly on areas of movement and not in static tissues.

## Conclusion

Using deformation fields, a revisited conjugate-gradient descent with exact line-search, a new density compensation and a new coil-sensitivity estimation, we have developed a new approach to compressed sensing image reconstruction for ferumoxytol-enhanced radial free-running cardiac image acquisitions. Indeed, objective and subjective image quality metrics suggest a potential for anatomical and functional image quality improvements on the one hand or hold promise for scan time abbreviations on the other. Clinically relevant parameters associated with cardiac anatomy and function are consistent with those extracted from contemporary reference standard methods.

## Data Availability

All data produced in the present study are available upon reasonable request to the authors.

## Supplementary Material

### Supplementary Methods

#### New density compensation

The sampling trajectory of the free-running sequence used for all acquisitions in this study was a radial trajectory, where the sequence of radial lines follows a 3-dimensional (3D) phylotaxis pattern (42). In particular, each line crosses the center of the Fourier space, so that the trajectory covers a ball centered in **0**. Let us index the complete list of radial lines with indices 1, …, *nLine*_*tot* where “*nLine*” means “number of lines” and “*tot*” refers to the complete set of acquired lines of the free-running sequence. Let be ***p***_***i***_ the location in *k*-space of the starting point of radial line number *i*. Then is

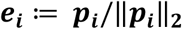

the only unit vector pointing in the direction of ***p***_***i***_. We will admit the presence of a spatial laboratory frame with orthonormal basis vectors ***e***_***x***_, ***e***_***y***_, ***e***_***z***_. The unit sphere is intersected by the x-y-plane on a circle that we will call the “equator”, which disconnects two open unit half spheres: one containing points with positive z-coordinates (open upper half sphere) and another one containing points with negative z-coordinates (lower open half sphere). We will call “closed upper half sphere” the union of the equator and the open upper half sphere, which is in fact a closed set. The closed lower half sphere is defined accordingly.

For simplicity, and without limiting generality, we will admit that ***e***_**1**_, …, ***e***_***nLine***_***tot***_ are contained in the closed upper half sphere. The complete list ***e***_**1**_, …, ***e***_***nLine***_***tot***_ samples the closed upper half sphere quite uniformly in practice. However, the binning operation (described in the reconstruction section) selects a subset of lines for each data bin. Let us call *nLine* the number of lines in a given bin and let be {*i*_1_, …, *i*_*nLine*_} ⊂ {1, …, *nLine*_*tot*} the index subset of lines included in that bin. Therefore, the list 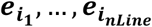 do not longer samples the previous half sphere uniformly. It follows that the trajectory points in that bin, although uniformly distributed radially, are not uniformly distributed spherically.

The reconstruction requires an estimation of the finite volume elements for each point of the *k*-space trajectory, the inverse of which is known as “density compensation”. The more natural estimation of these volume elements is achieved by computing the volume of the Voronoi cell for each trajectory point. The number of points being in practice so large, it is unpractical to perform the Voronoi algorithm of a concrete list of trajectory point in 3D space. Instead, we propose here to estimate those 3D volume elements from an estimate of the solid angles of the 2D spherical Voronoi cells of points 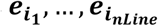 on the unit sphere. (See supplementary figure S2)

Given any list of points 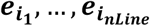 of the unit sphere, the 2D spherical Voronoi cell of point 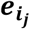 is by definition the set of points on the unit sphere that are closer to 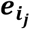 than to any other of the list. Of course, we consider that the distance between two points on the unit sphere is the length of the geodesic between them. Each such Voronoi cell is a 2D spherical surface elements which covers a solid angle. In order to estimate that solid angle with the 2D Voronoi algorithm, we perform a projection of 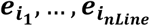 to the plane parallel to x-y-plane with z-coordinate equal to 1 in a similar (but different) way to stereographic projection: we simply rescale each unit vector so that its z-component is equal to 1. The 2D Voronoi algorithm is then performed on these projected points (see supplementary figure S2). The surface of each 2D flat Voronoi cell is assigned back to the corresponding point on the sphere by multiplying it with a correction factor that depends on the polar angle *θ* and which is given by cos^3^(*θ*), and leading this way to an estimate of the solid angle of each 2D spherical Voronoi cell. That process is repeated for points on the closed lower half sphere by projecting points on a plane with z-coordinate equal to −1. The points close to the equator are however projected quite far away as compared to most of the other points and the error on their 2D spherical surface estimate is relatively large. Moreover, the Voronoi algorithm assigns a non-sense value to point located in the periphery of the set of points. For that reason, the Voronoi cells for points located close to the equator are estimated separately by repeating the entire process with different half-spheres: once with the two half-sphere disconnected by the x-z-plane, and once with the two half-spheres disconnected by the y-z-plane. The 2D Voronoi algorithm is thus ran six times in total in order to cover the entire sphere.

Finally, the volume element of any point on the 3D radial trajectory is estimated as follows. Let be ***k*** a trajectory point on a radial line. Let be Δ*r*^*next*^ the distance to its next neighbor on the same line, and let be Δ*r*^*prev*^ the distance to its previous neighbor on the same line. We average them to obtain the distance Δ*r*. If ***k*** has only a next neightboor we set Δ*r* = Δ*r*^*next*^ and we set Δ*r* = Δ*r*^*prev*^ in the other case. Let be Δ*S* the solid angle assigned to the radial line supporting ***k***. Then we set the volume element of ***k*** equal to 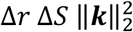. There is an exception for the *k*-space center ***k*** = 0. In that case we compute an average radius *r* as the average half distance to all its immediate neighbors. We set the its volume elements equal 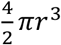.

#### New coil-sensitivity estimation

The coil-sensitivity estimation presented here is a modification of the method presented in (53). The difference is that we don’t use polynomial fitting. In this subsection, we will call Ω the set of points in space (or voxels, or pixels) where protons are sufficiently abundant to generate a signal amplitude significantly larger than noise amplitude, while Ω^*c*^ (the complement set of Ω) will be the set of space locations where only noise is observable on images, such that the air around the patient and some parts of the lungs. We will call *C* the coil-sensitivity, which is a spatially dependent complex value.

To estimate *C* in Ω, we made use of the low-resolution “prescan” images acquired automatically as part of the sequence protocol. These images are acquired sequentially with the exact same parameters for both the array-coils (or surface coils) and with the two body-coils located around the tunnel of the scanner. A reference anatomical image was constructed as the root-mean-square image of the two body-coil images (discarding thus the phase information). Thresholding this image with a visually determined value allowed to build an image mask defining Ω and Ω^*c*^. Some remaining unwanted voxels in Ω (especially in the corners of the image) could also be removed manually or by intersecting Ω with a box-shape mask excluding the borders of the image. The coil-image of each element of the coil-array was then point wise divided by the reference anatomical image, resulting in one complex-coil sensitivity estimation for each coil. The phase of that coil-sensitivity estimation was thus the ones of the coil-images. We omitted to subtract the phase of one of the body-coil image in order to avoid singularities in the final coil-sensitivity estimation. Until here, our method is the practically the same as presented in (53). But that former method uses polynomial fitting for two purposes: to reduce the noise in the coil-sensitivity estimation in Ω and to extrapolate the coil-sensitivity in Ω^*c*^. In our method, the noise affecting *C* in Ω was filtered out to some extent by averaging the value in each voxel with the neighboring voxel values, but only considering the neighbors lying inside Ω. The average was normalized according to the number of neighbors for each voxel and the averaging process was repeated iteratively until satisfactory result. In order to perform the extrapolation of *C* in Ω^*c*^, we solved the following Laplace boundary value problem in Ω^*c*^ for the real part of *C* that we will call *rC* :

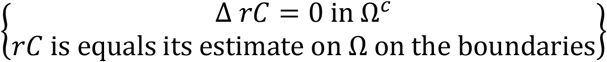

and we did exactly the same for the imaginary part *iC*. Circular boundary condition where enforced on the border of the image and the boundary value problem was solved with the well known Jacobi algorithm for Laplace equation. The complex coil-sensitivity estimate was then formed as *C* = *rC* + *j iC* where 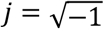.

In the study 2, we compare the above described coil-sensitivity to three others, among which two make use of polynomial fitting (one for extrapolation and one for both noise reduction and extrapolation).

#### New implementation of motion-compensated temporal-total-variation (MC-tTV) for 3D images

While MC-tTV for 2D images have been implemented for the MASTER reconstruction, we have implemented it for 3D images for our reconstruction. The mathematical formalism to describe MC-tTV is in principle independent of the image dimensionality and can therefore already be found in the literature. The innovation of our work, concerning MC-tTV, resides therefore in our 3D implementation and not in the concepts. The reason why we give here our view of the mathematical formalism describing MC-tTV is for giving indication on our implementation.

In the following, any MRI image ***x*** will be represented as a vertical concatenation ***x*** = [***x***^(1)^; … ; ***x***^(*nFr*)^] of 3D frames, the 3D frame ***x***^(*i*)^ being the static 3D image written as a column vector by column-major order and *nFr* being the total number of frames. The dynamic image ***x*** is thus a vector consisting of the concatenation of ***x***^(1)^ to ***x***^(*nFr*)^. *X* and *X*^(*i*)^ will be the vector spaces in which ***x*** and ***x***^(*i*)^are lying.

We define the temporal-total-variation (tTV) as

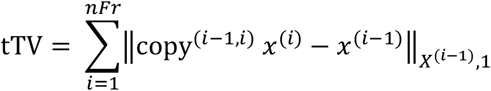

Here is copy^(*i*−1,*i*)^ a mathematical formality. It is the map that transports *x*^(*i*)^ ∈ *X*^(*i*)^ onto its identical copy in *X*^(*i*−1)^ so that we can subtract *x*^(*i*−1)^ from it and evaluate on it the 1-norm defined on *X*^(*i*−1)^. The vector copy^(*i,i*−1)^ *x*^(*i*)^ is component wise equal to vector *x*^(*i*)^ but interpreted as a vector lying in *X*^(*i*−1)^. We have thus

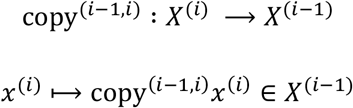

If we admit that it is clear that *x*^(*i*)^ is meant to lie in *X*^(*i*−1)^ when written inside the 1-norm 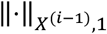, we can simply write

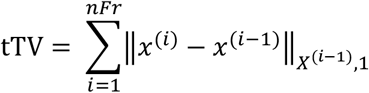

The one-norm 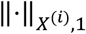 of a frame *w* ∈ *X*^(*i*)^ is defined as

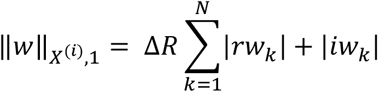

where *N* is the total number of voxels in the frame, Δ*R* is the voxel volume, *rw*_*k*_ is the real part of the voxel value of voxel number *k* in *w*, and *iw*_*k*_ is the imaginary part of the voxel value of voxel number *k* in *w* (here stands letter *i* in *iw*_*k*_ for “imaginary part”, it is not an index). We consider throughout the entire article that index *i* = 0 is identified with *i* = *nFr* and *i* = 1 is identified with *i* = *nFr* + 1, in a circular manner.

As explained in the reconstruction section, a first reconstruction is performed with tTV regularization only, leading to a list of frames *x*^(1)^, …, *x*^(*nFr*)^. For *i* = 1, …, *nFr*, an image registration between moving image *x*^(*i*)^ and reference image *x*^(*i*−1)^ was then performed in order to estimate the deformation-field *DF*^(*i*)^ defined as the local spatial translation

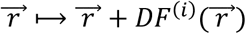

being so that *x*^(*i*)^ is deformed to match *x*^(*i*−1)^ as

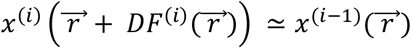

This registration was repeated independently for *i* = 1, …, *nFr* with the Niftyreg program (44,45) using default parameters (excepted maximum number of iterations): maximum number of iterations (1000), number of pyramidal levels (3), weight of bending penalty term (0.005), weight of L2-norm displacement penalty term (0.0), weight of linear elasticity penalty term ([0.0, 0.0]), final grid spacing (5 voxels), similarity measure (normalized mutual information with 64 bins), mask (no mask), type of deformation (spline).

Each deformation-field *DF*^(*i*)^ was then encoded into a deformation (sparse) matrix *T*^(*i*)^ which is so that

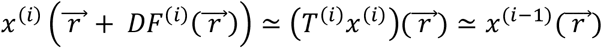

Or more concisely

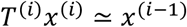

The map *T*^(*i*)^ is thus a linear map

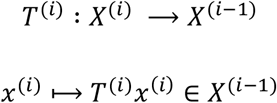

We define the motion-compensated tTV (MC-tTV) as

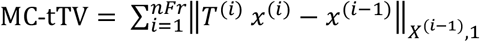

which is the sum of motion-compensated residuals between adjacent frames. The sparse matrix *T*^(*i*)^ was constructed as follows. Let be 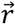 a voxel-center position and let be

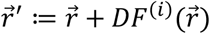

the new position given by the deformation-field in 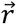. Position 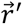 is not on the Cartesian grid in general. Let be 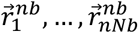 the immediate Cartesian neighbors of 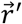 (i.e. the corners of the box containing 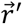) where “nb” stands for “neighbor” and “nNb” stands for “number of neighbors”. We define our (linear) interpolation of frame *x*^(*i*)^ at position 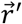 as the weighted sum

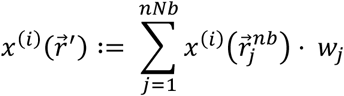

where the interpolation weights *w*_1_, …, *w*_*nNb*_ are normalized and given by

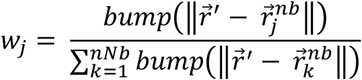

and where the *bump*-function is parametrized so, that it is non-zero if its argument is smaller than the voxel size and zeros else. In that way depends 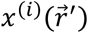 linearly on the immediate Cartesian neighbor of 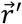. In the case where 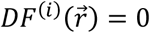 is 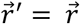 on the cartesian grid and 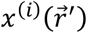 equals 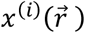. The link between a deformed frame and the frame itself is thus linear and the matrix of that linear map is the matrix we called *T*^(*i*)^. Given a deformation-field, the interpolation weights were calculated for all voxels and stored as coefficients of the sparse matrix *T*^(*i*)^ so that

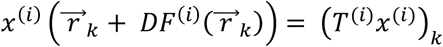

The transpose 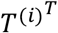 of sparse matrix *T*^(*i*)^, which is used during the iterative reconstruction, could then be evaluated using Matlab the build-in functions. A CSR sparse matrix format was developed in Matlab to store any sparse matrix (such as *T*^(*i*)^ and 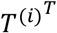) and an efficient matrix-vector multiplication was coded in C++ and parallelized with openMP. That code was then compiled as a Matlab executable file (MEX file) to allow the function call from any Matlab code.

#### Revisited Conjugate-Gradient-Descent (CGD) with exact line search

We summarize here some elements presented in details in an online freely available document (54). We make use of the conjugate-gradient-descent (CGD) as described in the original article (35), which presents two version (and some variants) of the algorithm. We make use of the second version, which is suited for solving the normal equation

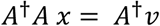

who’s solution set is equal to the set of minimizer of the least-square problem

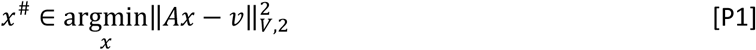

Here is *A* a linear map from vector space *X* to vector space *V*:

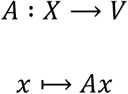

and *A*^†^ is the adjoint of *A*. Vector *x* ∈ *X* is the MRI image and vector *v* ∈ *V* is some given (constant) vector. The 2-norm ‖⋅‖_*V*,2_ on space *V* is induced by an Euclidean product as

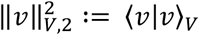

As explained in (54), this Euclidean product ⟨⋅ | ⋅⟩_*V*_ is given by

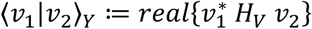

where *H*_*V*_ is a Hermitian positive definite matrix related to the list of volume elements (inverse density compensation) of the k-space trajectory points and also to the voxel volume (see next subsection for concrete definition).

On vector space *X* we use another 2-norm ‖⋅‖_*X*,2_ given by

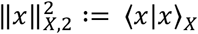

where the Euclidean product ⟨⋅ | ⋅⟩_*X*_ is given by

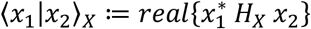

where *H*_*X*_ is a Hermitian positive definite matrix equal to the voxel volume Δ*R* time the identity on *X*.

The adjoint *A*^†^ of homomorphism *A* is the only one that verifies

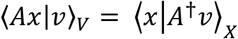

for any *x* ∈ *X* and any *v* ∈ *V* and is given in term of matrices by

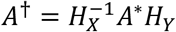

where matrix *A*^∗^ is the conjugate transpose of matrix *A*.

In order to perform the CGD algorithm to solve problem P1, one must perform the CGD algorithm as given in (35) but with the adjoint matrix *A*^†^ instead of the conjugate transpose *A*^∗^ because our Euclidean products are not standard (i.e. the matrices defining the Euclidean products are different from the identity).

In our reconstruction, the least square problem we solve with CGD is the lest-square sub-problem of ADMM given by

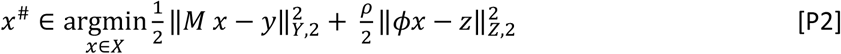

where *M* = *FC* is the model, in our case the composition of the coil-sensitivity map and the (possibly non-uniform) Fourier transform, and *ϕ* is the sparsifier transform. The variable *z* is one of the other decision variable of ADMM algorithm described later. The data vector *y* (which is the raw-data measured by the MRI machine) stands in a vector space *Y* which is endowed with ist own Euclidean product given by

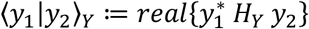

where *H*_*Y*_ is a Hermitian positive-definite matrix defined concretely in the next sub-section. Vector *z* stands in another vector space *Z* which is also endowed with ist own Euclidean product given by

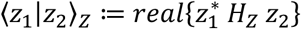

where *H*_*Z*_ is also a Hermitian positive-definite matrix defined concretely in the next sub-section. The map *M* is from *X* to *Y* while the map *ϕ* is from *X* to *Z*. The 2-norms on *Y* and *Z* are naturally given by

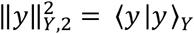

and

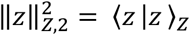

Finally, the real parameter *ϱ* is a positive parameter of the ADMM algorithm. In order to solve P2 with the CGD, it must be rewritten as a least square problem with one term. We do it as follows.

We define the map *A* by the matrix

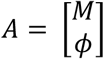

which is a map from *X* to *V* ≔ *Y* × *Z*. We define

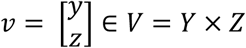

On the vector space *V* we define the Euclidean product

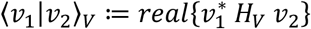

where *H*_*V*_ is the block matrix

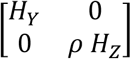

One can check that given these definitions, it holds

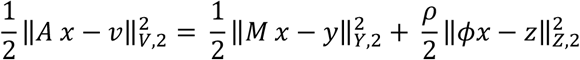

The lest-square sub-problem of ADMM algorithm can then be solved by solving the least-square problem

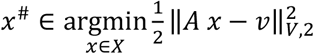

where *A* is the linear map, *v* is the constant data and *x* is the decision variable, which is nothing else than problem P1. Solving this problem with CGD implies to make use of *A*^†^, the adjoint operator of *A*. It is a consequence of all definitions above that it holds

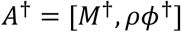

We give in (54) a recipe to solve, with CGD, least-square problems such as problem (P1) and (P2).

The original CGD algorithm presented in (35) makes use of standard Euclidean product on *X* and *V* and the adjoint of *A* is in that case given by the complex-conjugate transpose matrix *A*^∗^. The use of non-standard Euclidean products ⟨⋅ | ⋅⟩_*X*_ and ⟨⋅ | ⋅⟩_*V*_ has the consequence that the adjoint *A*^†^has to be used instead of *A*^∗^. This allows to perform the CGD algorithm in a proper way with exact line search. In fact, the line-search parameter *a* can be evaluated properly as

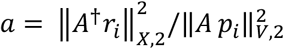

where *r*_*i*_ is the current residual and *p*_*i*_ is the current descent direction. We see in the previous equation that the adjoint *A*^†^ is used (and not the conjugate-transpose *A*^∗^) and that the non-standard 2-norms 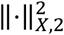 and 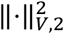 play a role. The image can then be updated as

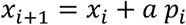

We refer the reader to (54) for detailed information.

We do not pretend to publish a “new conjugate-gradient-descent” in the present article but the way we implemented it, taking into account the different inner product matrices *H*_*X*_, *H*_*Y*_ and *H*_*Z*_, is new for MRI reconstruction as far as the authors knows.

#### The compressed sensing reconstruction with MC-tTV regularization

We define 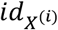 to be the identity map on vector space *X*^(*i*)^. As defined above, *T*^(*i*)^ is the map that transform frame *x*^(*i*)^ into a frame in *X*^(*i*−1)^ that resembles frame *x*^(*i*−1)^. We define the linear map *ϕ* by its matrix as

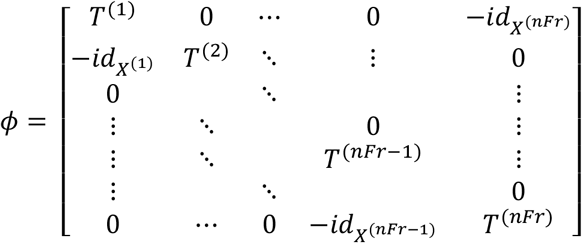

By the definition of the maps *T*^(*i*)^ and 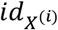, the map *ϕ* is from vector space

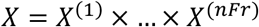

to another vector space that we will call *Z* and which is defined by

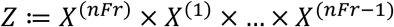

It holds then

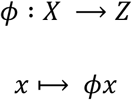

Vector space *Z* is of course identical to vector space *X* since every *X*^(*i*)^ is a copy of ℂ^*N*^ with *N* being the number of voxels in each frame. But the use of different labels allows to highlight in which order the frames are written, which is different in *X* and *Z*. One can check that multiplying the vector

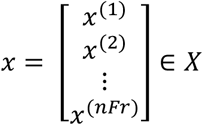

by matrix *ϕ* leads to

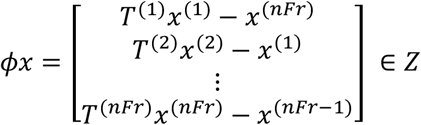

Given the 1-norm 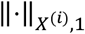 as defined in the subsection above, we define the 1-norm ‖⋅‖_*Z*,1_ on *Z* by

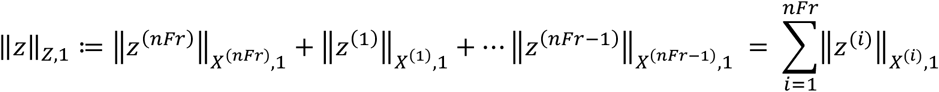

It follows

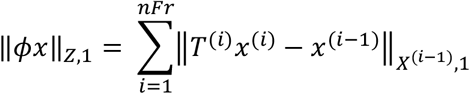

which is nothing else than MC-tTV. Of course, the index (*i* − 1) is to be interpreted in a circular manner in the previous sum.

As defined above, *M* = *FC* is the model that assigned a data vector *FCx* to each image *x*. Vector *y* is the vertical concatenation of the raw-data bins *y*^(1)^, …, *y*^(*nFr*)^. Each vector *y*^(*i*)^ is a column vector lying in 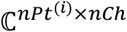 where *nPt*^(*i*)^ is the number of trajectory points in bin number (*i*) and *nCh* is the number of channels (or coils) used for the acquisition. We define 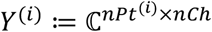 to be the vector space that contains *y*^(*i*)^ and we define the data-space

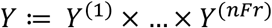

which is the vector space that contains *y*. On vector space *Y*^(*i*)^ we define the Euclidean product 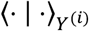 by

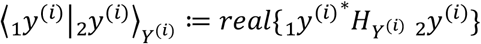

where the star symbol ⋅^∗^ stand for the complex conjugated transpose and 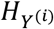 is the diagonal matrix

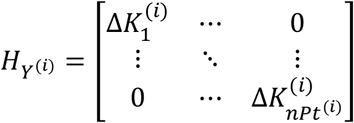

where 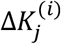 is the volume element in k-space (i.e. inverse density compensation) for point number *j* in the trajectory of bin number *i*. This induce the Euclidean product ⟨⋅ | ⋅⟩_*Y*_ on *Y* given by

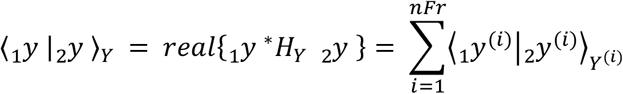

where *H*_*Y*_ is the diagonal block matrix

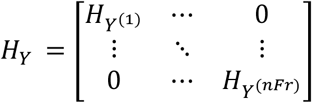

This Euclidean product on *Y* induces the 2-norm 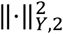 given by

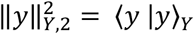

On vector space *Z* we define the Euclidean product as on vector space *X* :

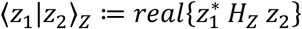

where *H*_*Z*_ is a Hermitian positive definite matrix equal to the voxel volume Δ*R* time the identity on *Z*. The definitions are so that both *H*_*X*_ and *H*_*Z*_ can be replaced by the scalar Δ*R*. The 2-norm on *Z* is then given by

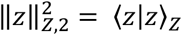

and one can easily check that

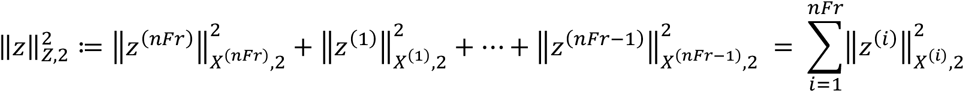

We have so far define the 2-norms on *X, Y* and *Z* as well as the 1-norm on *Z* and we are now ready to formulate the optimization problem of the reconstruction as a least-square problem regularized by MC-tTV:

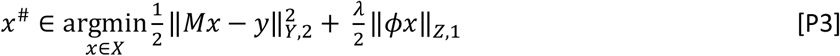

Problem (P3) is a generalized-LASSO problem and can be solved with the ADMM algorithm. The constant *λ* is a regularization constant. The ADMM algorithm for solving (P3) can be found in (43). We formulate the algorithm for our present need as follows (54):

##### INITIALIZATION

a) Chose a real positive constant *ρ*.
b) Initialize the variables *x*_*curr*_, *z*_*curr*_ and *u*_*curr*_

##### DO

c) Solve 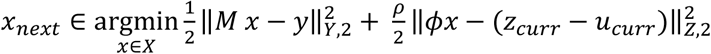
d) 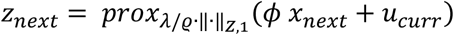
e) *u*_*next*_ = *ϕ x*_*next*_ + *u*_*curr*_ − *z*_*next*_
f) Update (*x*_*curr*_, *z*_*curr*_, *u*_*curr*_) ⟵ (*x*_*next*_, *z*_*next*_, *u*_*next*_)

### UNTIL some stopping-criterion is satisfied

Step (d) is straight forward and step (e) consists in applying the proximal operator associated to the norm ‖·‖_*Z*,1_ (see (54) for details) which is also straight forward. Step (c), in contrast, involves the solving of a least-square problem with two terms. To solve it with the CGD algorithm, it must be reformulated as a least-square problem with one single term as described above. Just set

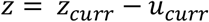

to recover P2. Again, we invite the reader to consult (54) for details.

### Ferumoxytol

The ferumoxytol contrast agent specifications are: Feraheme, polyglucose sorbitol carboxymethylether iron oxide, distributed by AMAG Pharmaceuticals, Inc., Waltham, MA02451.

### Study 2: Comparison of different coil-sensitivities using MASTER_3D+

All reconstructions in the present study 2 were performed using MASTER_3D+. Only the coil-sensitivity estimation was varied.

We name “coil-sensitivity A” the one explained above i.e., our new coil-sensitivity estimation presented in the section above.

We named “coil-sensitivity B” the coil-sensitivity obtained by using the Matlab function “adapt array 2d” (Ricardo Otazo, NYU) and implemented following (55) and (56). This method estimates the coil-sensitivity without use of the body-coil prescan images. It is the coil-sensitivity estimation used in XD-GRASP-5D_LAUS.

We named “coil-sensitivity C” the coil-sensitivity obtained by a method like method A, with the difference that a polynomial fit of degree 3 is used (for real an imaginary part independently) in order to extrapolate the coil-sensitivity in the volume without signal Ω^*c*^ (such as the air part in the lungs) The coil-sensitivity inside Ω remain therefore the same as coil-sensitivity A.

We named “coil-sensitivity D” the coil-sensitivity obtain by the method presented in the master thesis (57). This method could be applied to estimate the coil-sensitivity thanks to the MATLAB code present in the thesis itself. This method uses polynomial fitting for both reducing the noise of the coil-sensitivity estimate in Ω and for extrapolation in areas where coil-sensitivity cannot be measured (i.e. in Ω^*c*^).

We used three quantitative markers to compare the reconstruction performed with the four coil-sensitivity methods listed above. The first maker was the EF-bias as defined in study 1.

The second marker is a quantification of how spatially homogenous the reconstructed image was. This marker was so defined that a high spatial inhomogeneity of the image (typically a very bright signal close to the coils and a dark area far from the coils) resulted in a high value for the marker. This marker measures the standard-deviation of image values in the periphery of the thorax and was called “std-periphery” for that reason. It was constructed as follows. For each patient, an axial plane through the thorax in the reconstructed image was chosen. A mask was drawn manually around the periphery of the thorax on that transverse plane. We will call this mask *M*_*periph*_. Images reconstructed with coil-sensitivity A, B, C and D were then masked by *M*_*periph*_. The masked values were normalized by their average, separately for each coil-sensitivities. The standard deviation of the obtained list of values were then reported for each coil-sensitivity and each patient for statistical analysis. A list of standard deviation (running over patients) was thus obtained for each coil sensitivity. This was considered as a quantitative measure of the homogeneity of the image in the periphery of the thorax.

As a third quantitative marker of image-quality, the lung-liver interface sharpness (LLIS) was used. This was evaluated using the same method as the blood-muscle interface-sharpness used in study 1. For each patient, 6 points where manually chosen on the lung-liver interface on a coronal plane and the lung-liver interface–sharpness was evaluated on these points as described in (47). This sharpness was evaluated on the same coronal plane and the same 6 points for all four coil-sensitivities.

The significance of the average difference between any two lists of values was assessed with a paired student t-test. The difference was considered as significant for a p-vale smaller that 0.05.

### Study 3: Convergence analysis

To monitor the convergence of our new reconstruction (MASTER_3D+), the value of the data-fidelity term, of the MC-tTV and of the objective function were was saved at every iteration, together with 3 orthogonal image planes (transverse, coronal, and sagittal) of the first frame, which were called “monitor-images”.

The monitor images were visually assessed to confirm the convergence of the reconstruction, while the data-fidelity, MC-tTV, as well as the objective function were plotted graphically in order to confirm as good as possible a plausibly convergent scenario.

In absence of ground truth for in-vivo data, it was impossible to evaluate if the reconstructed images were fact converging to the ground-truth along the iterations. This was therefore assessed with simulated data obtained by sampling the k-space of synthetic images provided by the XCAT phantom (49–51). The same plots and monitor images were then displayed as for in-vivo data, and in addition, the sum-of-square error between ground-truth and reconstructed image along the iterations could also be observed.

### Additional reconstructions

Three additional reconstructions were performed on the data of patient 1 in order to give some elements of answers to some question about the registration strategies. We tested the reconstruction

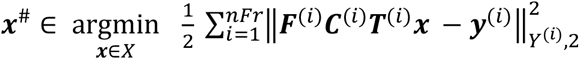

where a single static-frame ***x*** is reconstructed, and the deformation-field are part of the model: each ***T***^(***i***)^ deforms the static frame ***x*** on the frame ***x***^(***i***)^ corresponding to bin ***y***^(***i***)^. We named that reconstruction “**SENSE_with_motion_correction**”.

We also tested the reconstruction

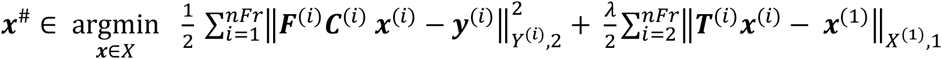

which is very similar to MASTER_3D+ but where the regularization term includes deformations from all frames to the first frame (all-to-one registration): each ***T***^(***i***)^ deforms the frame ***x***^(***i***)^ to match frame ***x***^(**1**)^. We named that reconstruction “**MASTER_3D+_all_to_first**”.

As another additional reconstruction, we re-estimated the deformation field on the result of MASTER_3D+, and we ran MASTER_3D+ again with these new deformation fields. We called the result **MASTER_3D+_bis**.

## Supplementary Results

### Result of Study 2: Comparison of different coil-sensitivities using MASTER_3D+

Table 5 displays all the results of study 2 and supplementary figure S5 does the same graphically. For the same reasons as in study 1, four patients were excluded from the EF measurements. The EF-bias was 0.0% ±1.9% for coil-sensitivity A (as given in study 1), it was −2.1% ±2.2% (p < 0.05) for coil-sensitivity B, it was 1.5 ±1.6 % (p < 0.05) for coil-sensitivity C and it was 1.4% ±1.3% (p < 0.05) for coil-sensitivity D. The EF-bias for coil-sensitivity A was not statistically different from 0, but the EF-bias for coil-sensitivity B, C and D was. EF measured with coil-sensitivities B, C and D were therefore significantly different from the reference 2D-CINE method. The marker “std-periphery” was significantly higher (more inhomogeneous spatially) for coil-sensitivity B than for any other method and there was no significant difference between other coil-sensitivity methods. The lung-liver interface-sharpness (LLIS) was significantly better for coil-sensitivities A and B than for coil-sensitivities C and D.

Supplementary figure S2 displays an axial plane of all coil-sensitivities (real part in top row and imaginary part in bottom row) that were used for patient 1. Supplementary figure S3 displays reconstructions of the same plane as supplementary figure S2 with the four different coil-sensitivities. Supplementary figure S4 display the same figure as supplementary figure S3 but masked with the mask that served to measure “std-periphery”.

### Result of Study 3: Convergence analysis

The monitor images showed a good convergence for all patient and no improvement was observed beyond 50 iterations, as far as visual inspection is concerned. Supplementary figure S6 shows an example of four selected monitor images in axial view. They were selected from MASTER_3D+ of patient 1 at iteration numbers 1, 10, 30, and 60. These images ensure that no major problem happened during the reconstruction and document that visual sharpness increases with iterations. Supplementary figure S7 shows the evolution of the data-fidelity, MC-tTV and objective-function as a function of the 60 iterations for MASTER_3D+ in patient 1. It provides an additional marker of convergence. Note that the decrease of the data-fidelity term is not monotonic, but there is no theoretical reason to be so.

Supplementary figure S8 displays the monitor images in axial plane of the reconstruction of the XCAT phantom data at iteration number 1, 10, 30 and 60. Sub-figure E displays the ground truth. Supplementary figure S9 displays the difference between the sub-images of supplementary figure S8 and the ground-truth. It demonstrates that the reconstructed image converges to the ground-truth almost everywhere.

Some error remains at the interface of different regions. Supplementary figure S10 displays the data-fidelity, the MC-tTV and the objective-function value along the 60 iterations of the reconstruction of the XCAT phantom data. Sub-figure D displays the evolution of the error (in term of L2-norm) between the reconstructed image and the ground-truth. All four graphs indicate convergence, especially sub-figure D which show that the error tends to zero.

### Results of additional reconstructions

Supplementary figure S11 shows, in coronal plane, the results on patient 1 of MASTER_3D+ (S11A) together with the additional reconstructions **SENSE_with_motion_correction** (S11C) and **MASTER_3D+_all_to_first** (S11D). Those two former reconstructions make use of “all-to-one” registrations, but supplementary figure S11 shows obviously that MASTER_3D+ (with adjacent frames registration) is superior. Moreover, supplementary figure S12 shows the registration residual (in one transverse plane) between adjacent frames (top row) and the corresponding registration residuals between the same frames and the first frame of the list (bottom row). Visual inspection shows that the first are sparser than the second, suggesting that the motion compensated residuals between adjacent frame is better for compressed sensing. Supplementary figure S11B shows, in coronal plane, the result of MASTER_3D+_bis, which is not superior to MASTER_3D+ as far as visual inspection can tell.

## Supplementary Discussion

The work presented in (29,58) for Cartesian acquisitions and in (16,59) for radial acquisitions is closely related to ours. These reconstructions have been achieved using deformation-fields to compensate for motion. But these techniques estimated the deformation between each frame and a single reference configuration (which was either a reference frame or a virtual configuration that do not appear among the frames). These all-to-one registration procedures did not take advantage of the small deformation amplitude between adjacent frames, which could potentially be a disadvantage compared to the registration between adjacent frames only. More comparison are of course needed in order to decide which registration strategy is better because the group-wise registration used in (58,59) had the advantage of being informed by all frames simultaneously. However, our preliminary experiments with SENSE_with_motion_correction and MASTER_3D+_all_to_first already suggest registrating temporally adjacent frames may be superior.

One major limitation of our reconstruction is that the deformation fields are not updated at every iteration of the reconstruction. We recognize that future developments should go into that direction. This could potentially accelerate the reconstruction because a second reconstruction informed by deformation fields would no longer be needed. But it is not certain that image quality would improve. The preliminary experiment with MASTER_3D+_bis did not show any significant improvement, as far as visual inspection can tell.

## Supplementary Figures

**Supplementary figure S1:**
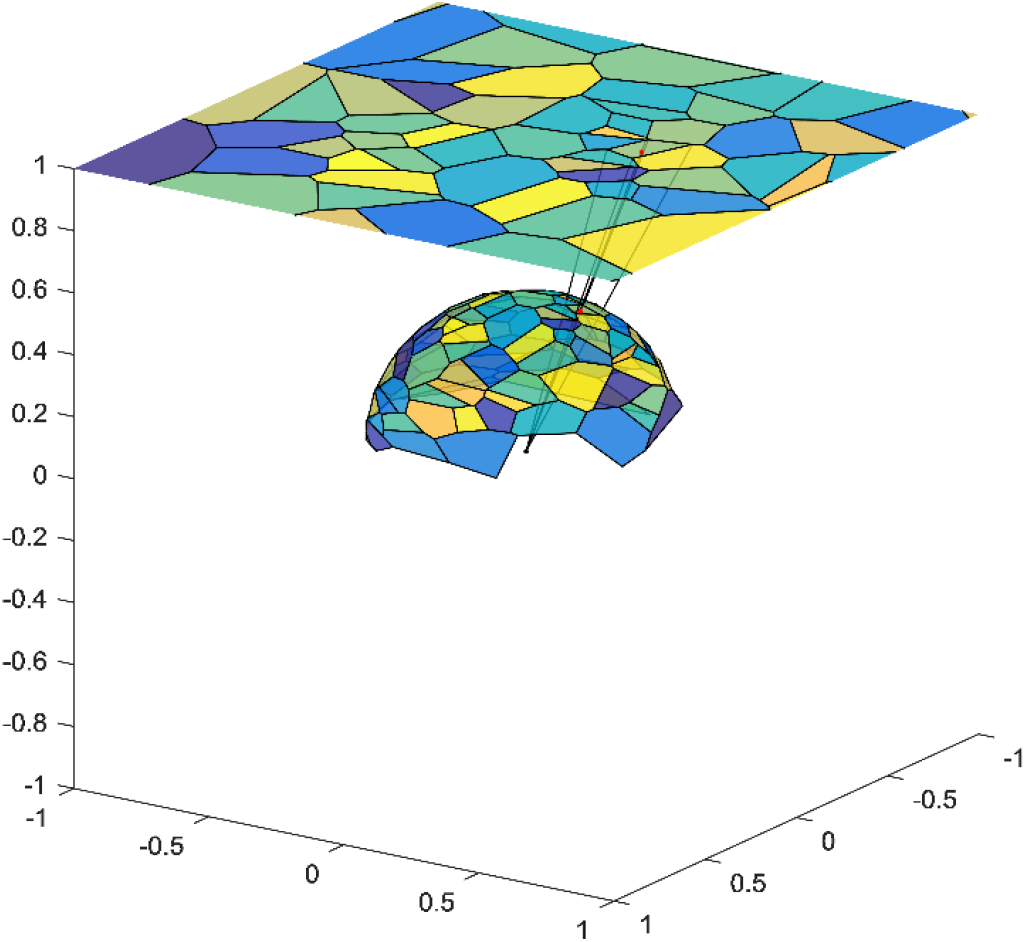
This picture describes de implementation of the spherical surface elements estimation by mean of the Voronoi algorithm performed on a 2-dimensional plane.

**Supplementary figure S2:**
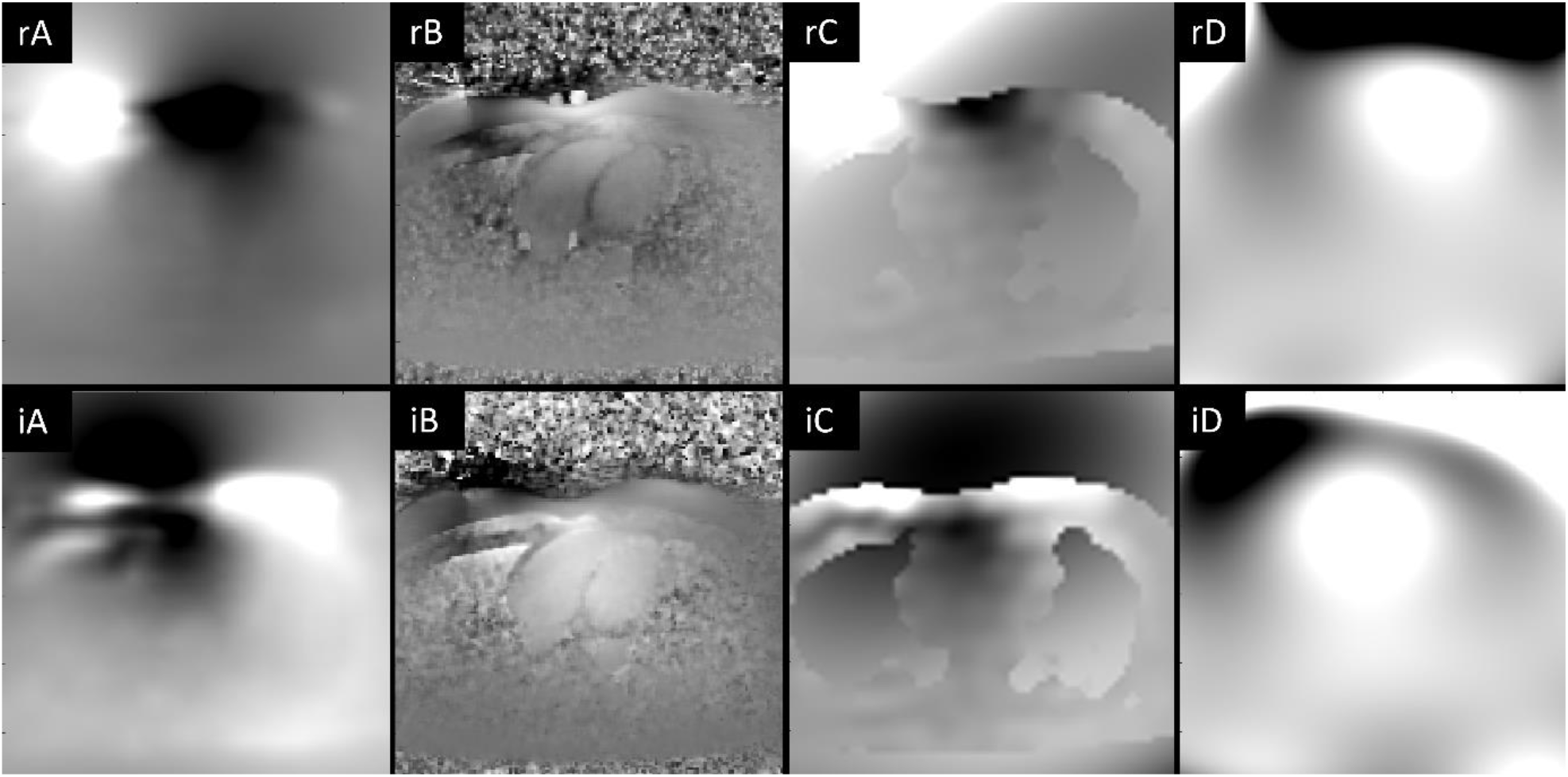
The sub-figures display the real-part (top row) of imaginary-part (bottom row) of the coil-sensitivity estimation performed with method A (left), method B (middle-left), method C (middle-right) and method D (right). “rA” stands for “real-part of coil-sensitivity method A”, “iA” stands for “imaginary-part of coil-sensitivity method A” and so on. Method A is our new coil-sensitivity estimation, while method B is the one used in XD-GRASP-5D_LAUS.

**Supplementary figure S3:**
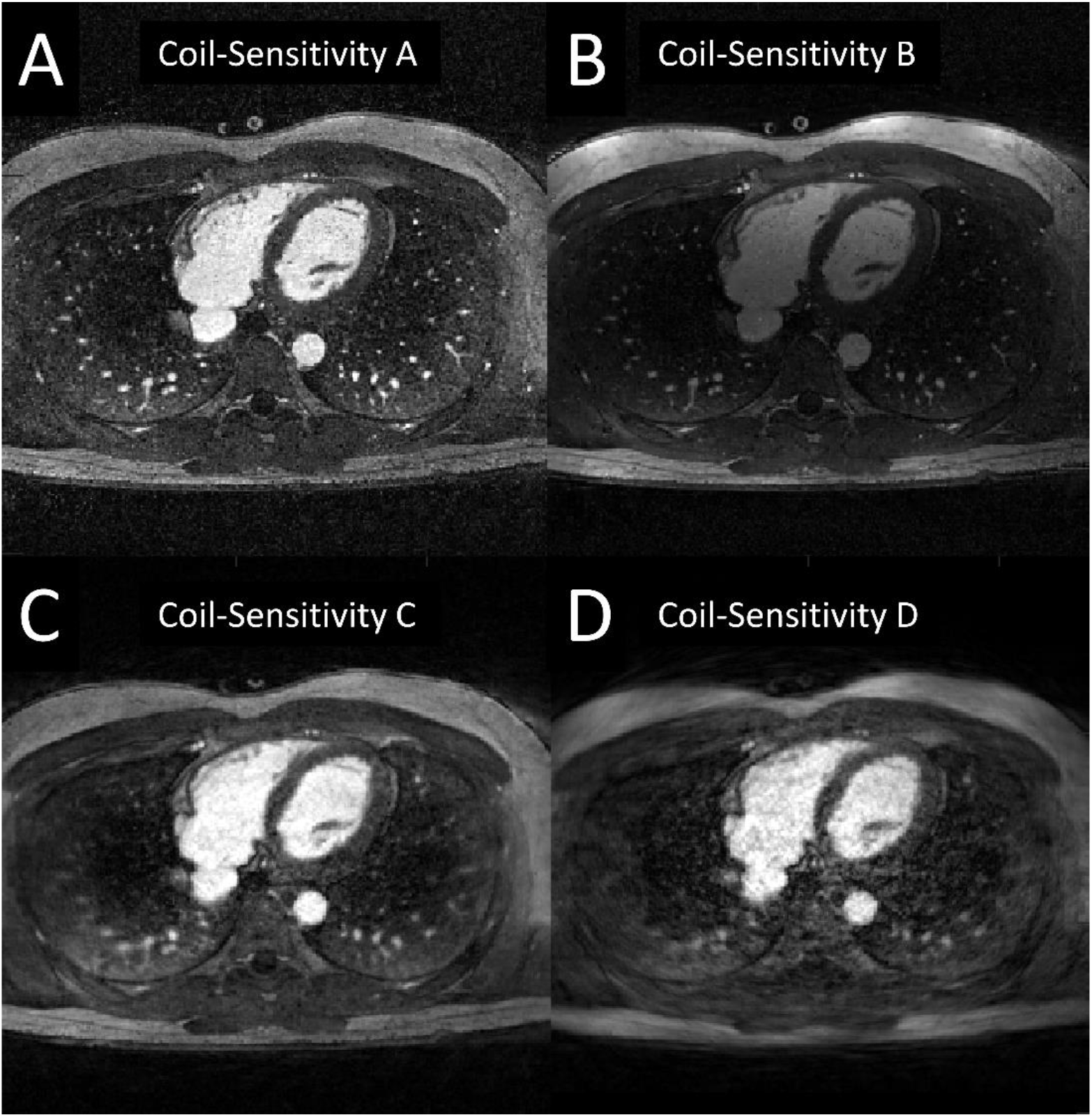
Each sub-figure displays the same plane for the same patient of the same reconstruction (MASTER_3D+) but using different coil-sensitivities. Sub-figures A displays the images reconstructed with coil-sensitivity A, and so on for coil-sensitivities B, C, and D. Method A is our new coil-sensitivity estimation, while method B is the one used in XD-GRASP-5D_LAUS.

**Supplementary figure S4:**
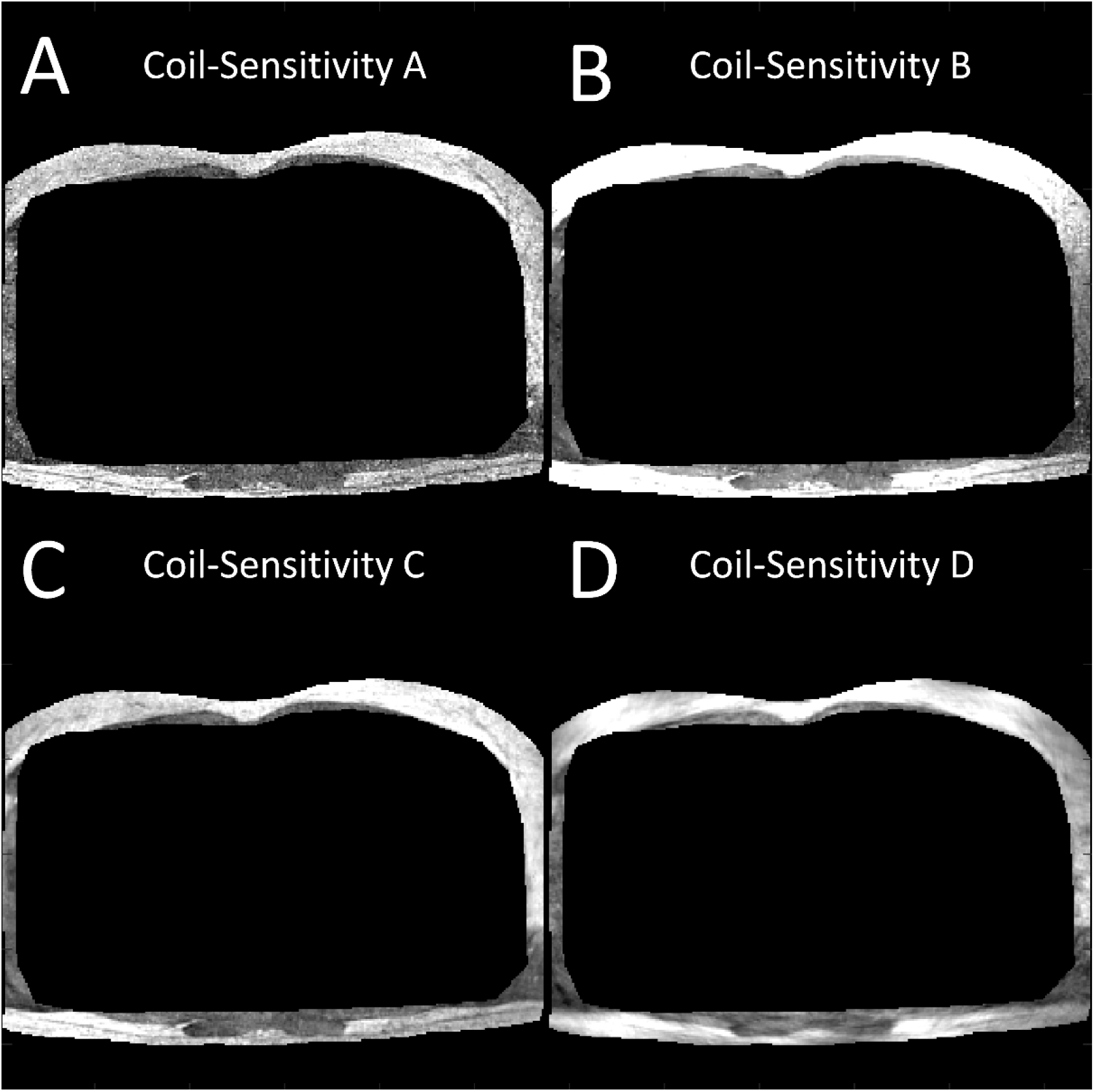
The same sub-figures as supplementary figure S3 are displayed but masked by the mask used for the measurement of the standard-deviation of peripheric values.

**Supplementary figure S5:**
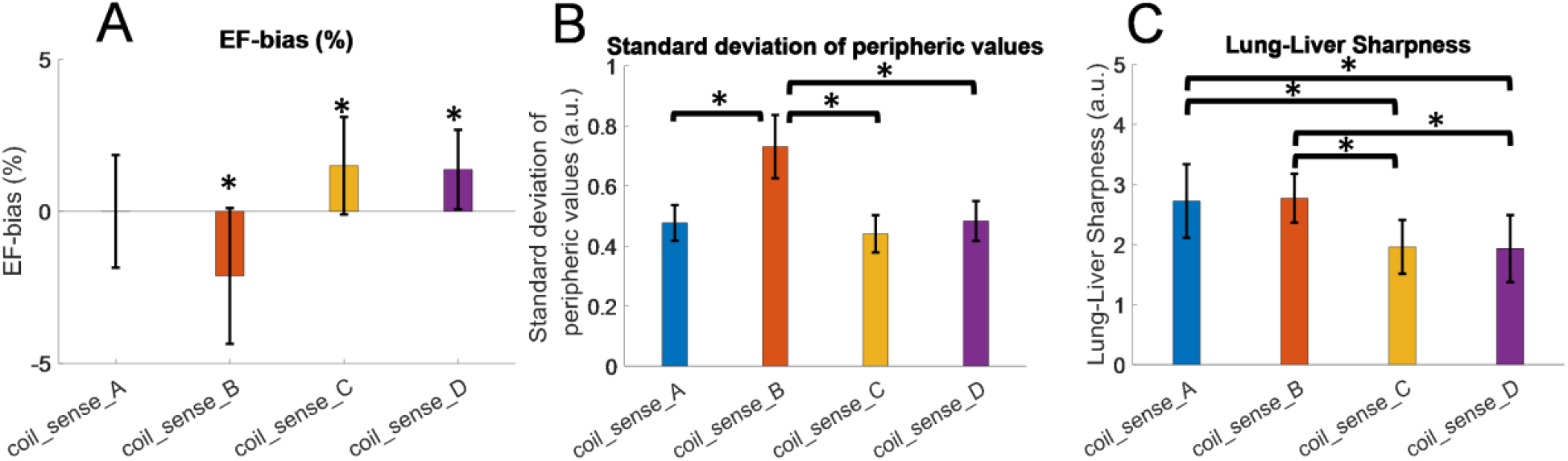
The results of supplementary table 5 are presented graphically in the present figure. The asterisc (*) in subfigure A designates an average significantly different from 0. The asteriscs (*) in sub-figures B and C designate an average difference significantly different from 0.

**Supplementary figure S6:**
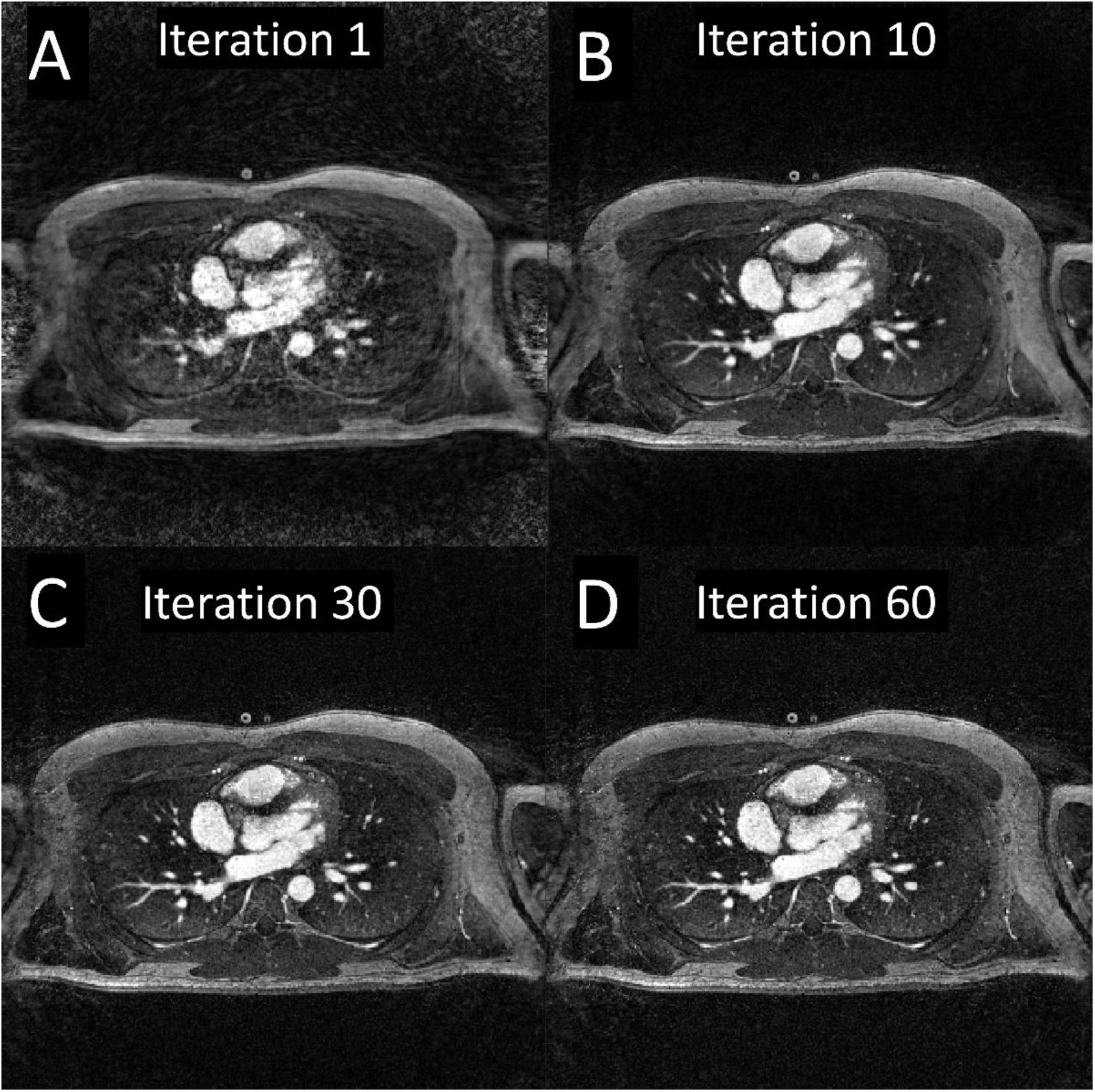
Selection of axial monitor-images of MASTER_3D+ of patient 1. The images were captured at iteration number 1 (A), 10 (B), 30 (C), and 60 (D) respectively.

**Supplementary figure S7:**
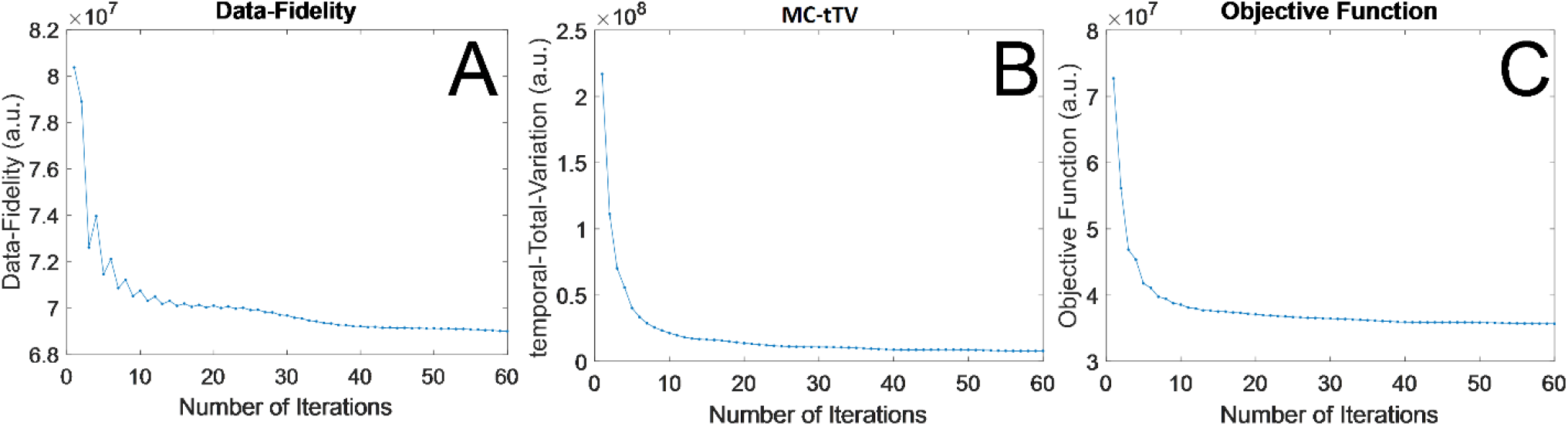
Example of the evolution of the data-fidelity term (A), motion-compensated temporal-Total-Variation (MC-tTV) (B) and objective function (C) along the 60 iterations for MASTER_3D+.

**Supplementary figure S8:**
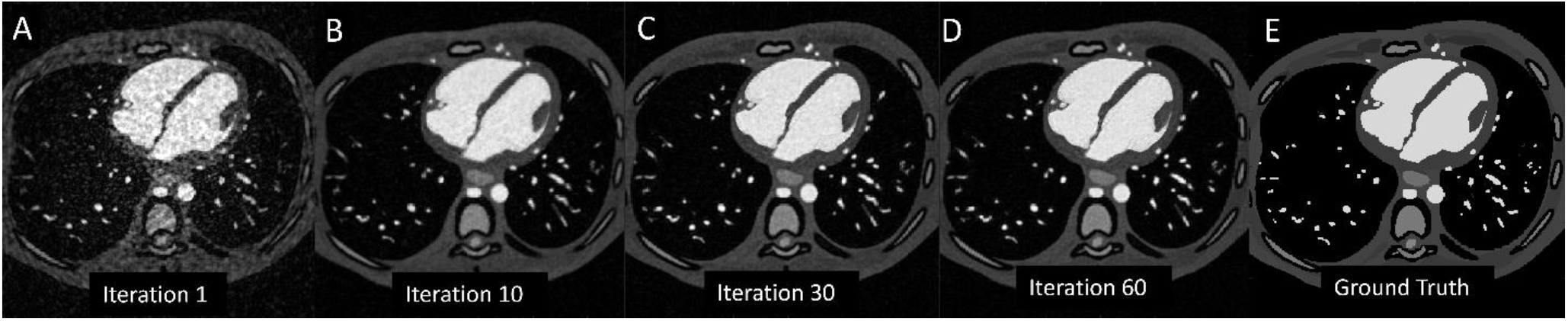
Selection of axial monitor-images of MASTER_3D+ for the XCAT numerical phantom. The images were captured at iteration number 1 (A), 10 (B), 30 (C), and 60 (D) respectively. Sub-figure E displays the ground-truth.

**Supplementary figure S9:**
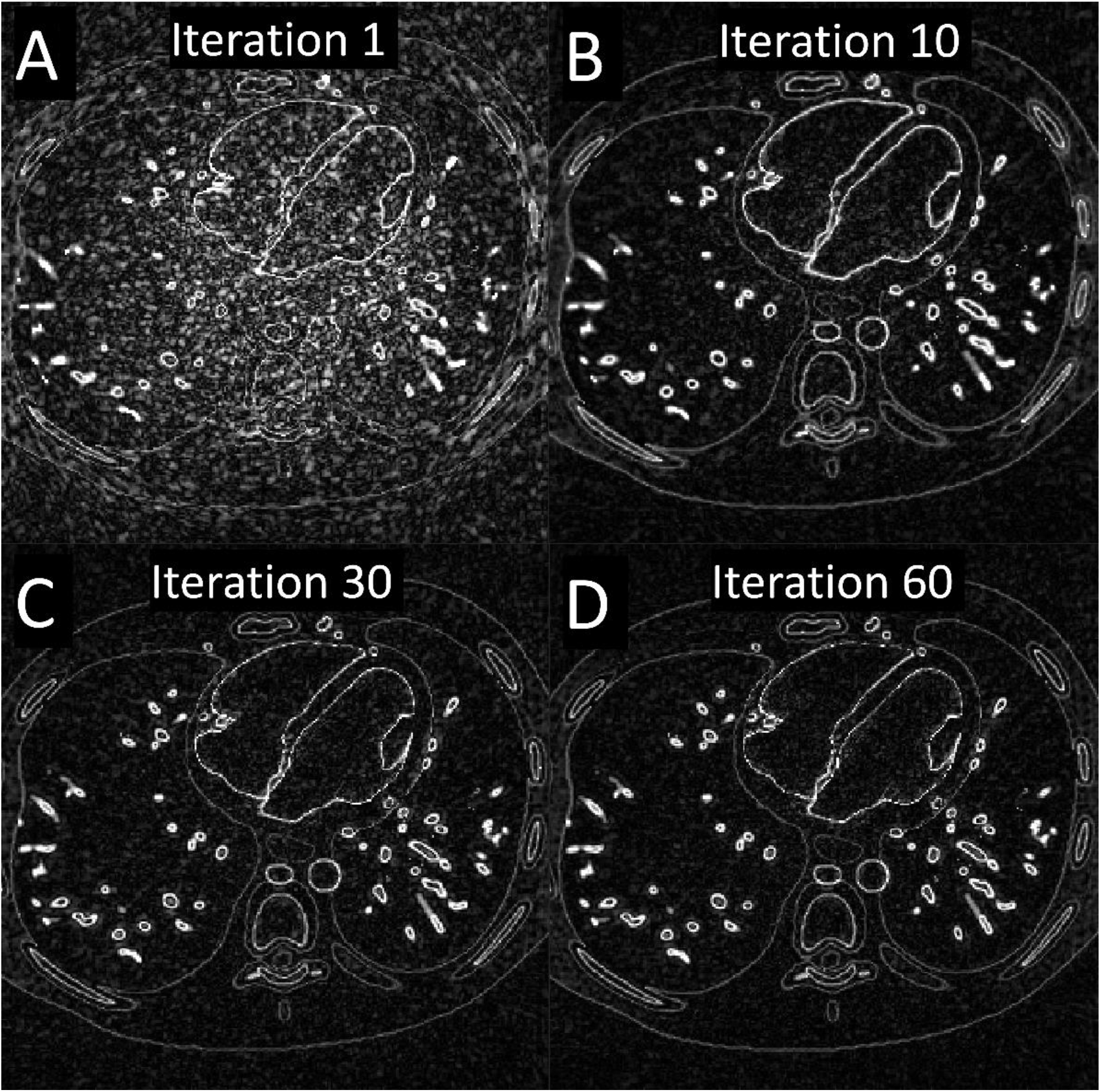
Difference between the MASTER_3D+ reconstruction of the XCAT phantom data and the ground-truth, at different iteration of the reconstruction: iteration number 1 (A), 10 (B), 30 (C), and 60 (D).

**Supplementary figure S10:**
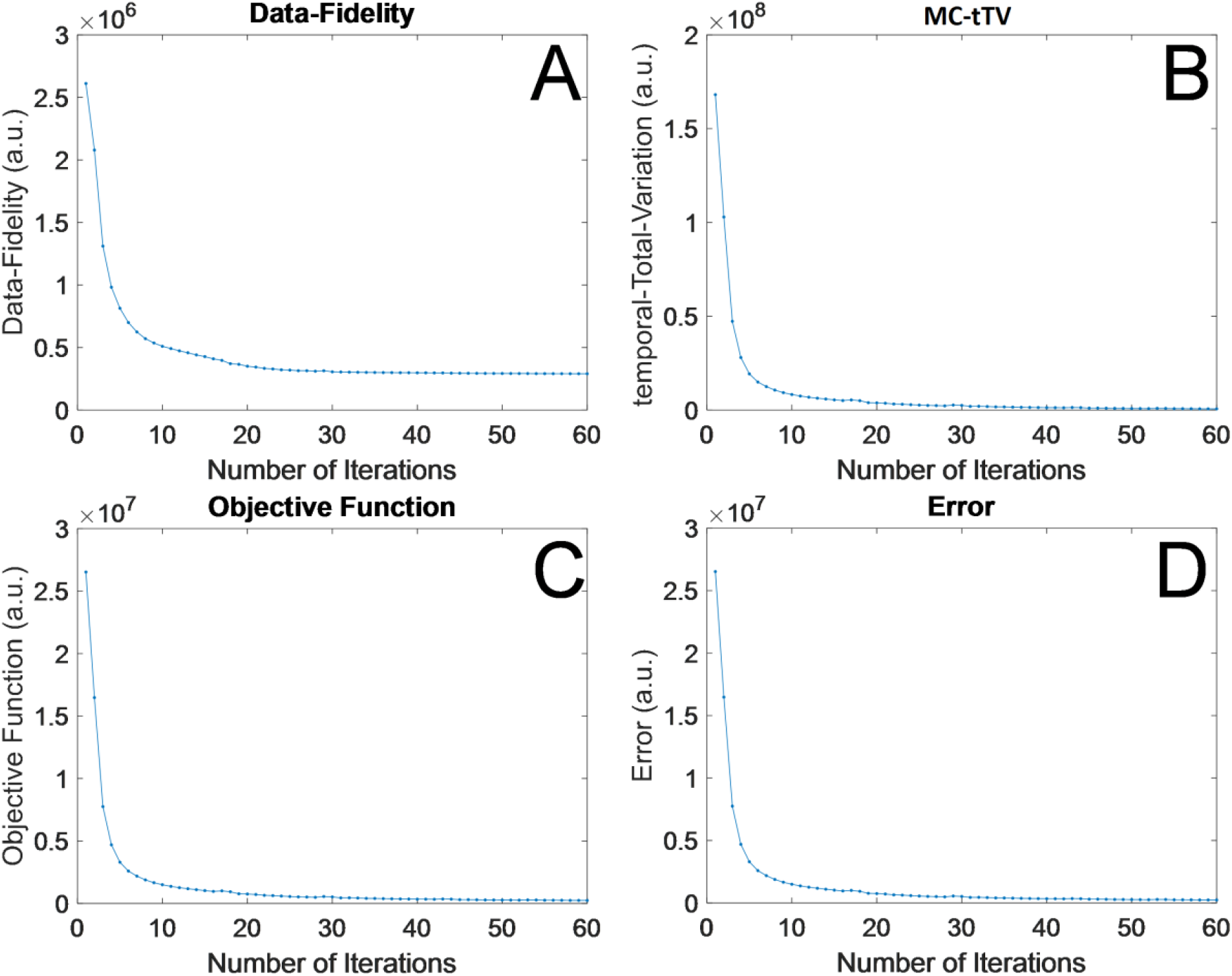
Evolution of the data-fidelity term (A), MC-tTV (B) and objective function (C) along the 60 iterations for MASTER_3D+ reconstruction of the XCAT phantom data. In contrast to reconstructions performed on patients, we can also plot here the error between the reconstructed image and the ground-truth at every iteration (D).

**Supplementary figure S11:**
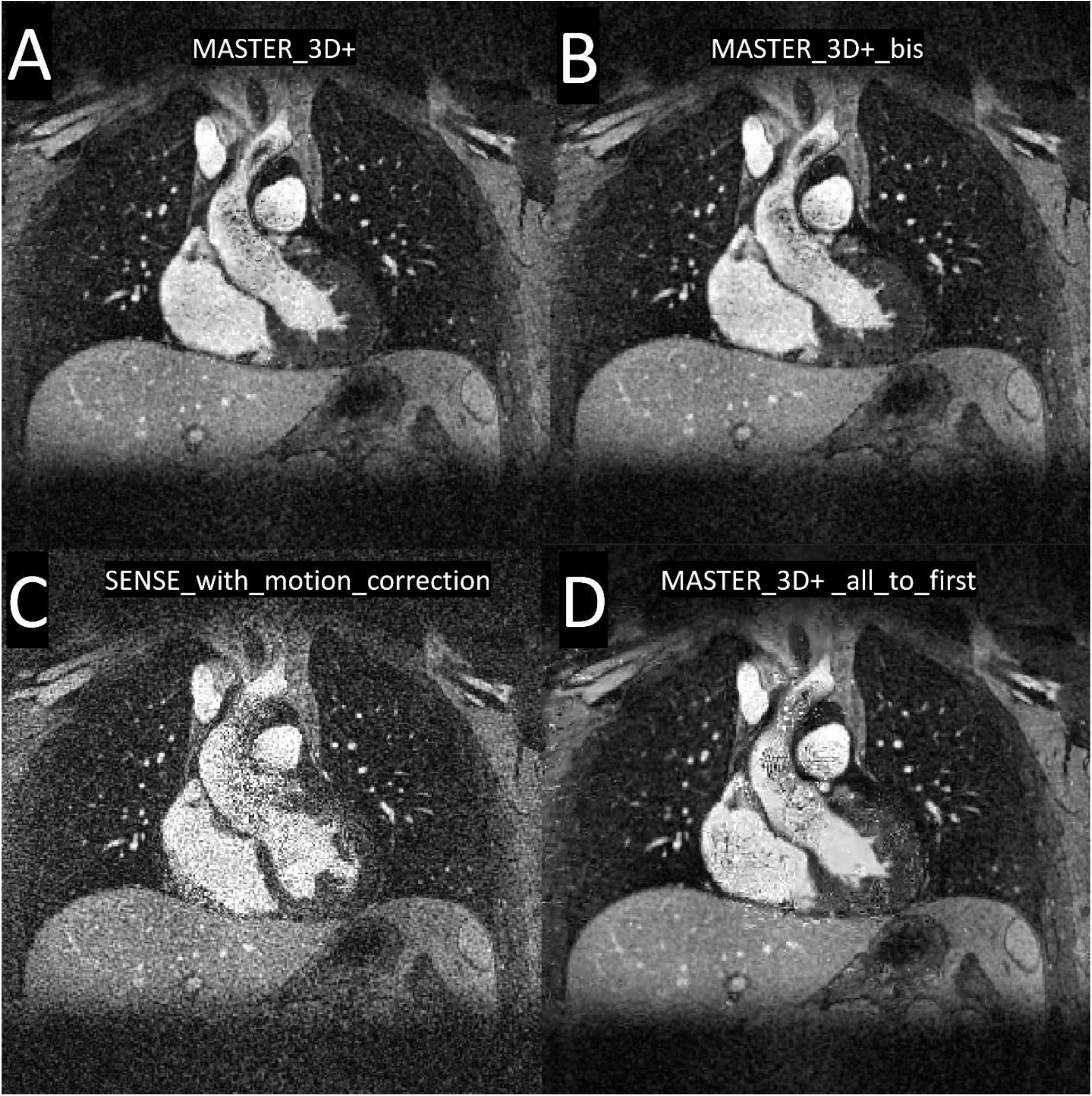
Sub-figure A displays MASTER_3D+ on patient 1. The deformation-fields were re-estimated on the result of MASTER_3D+ and the reconstruction was run again. The result is displayed in sub-figure B and is practically identical to the reconstruction displayed in sub-figure A. We called it “MASTER_3D+_bis”. Sub-figure C displays the reconstruction “SENSE_with_motion_correction” in which the deformations are incorporated inside the data-fidelity term and there is no regularization term. Sub-figure D displays the reconstruction “MASTER_3D+_all_to_first” where all frames are deformed to match the first frame in the regularization term.

**Supplementary figure S12:**
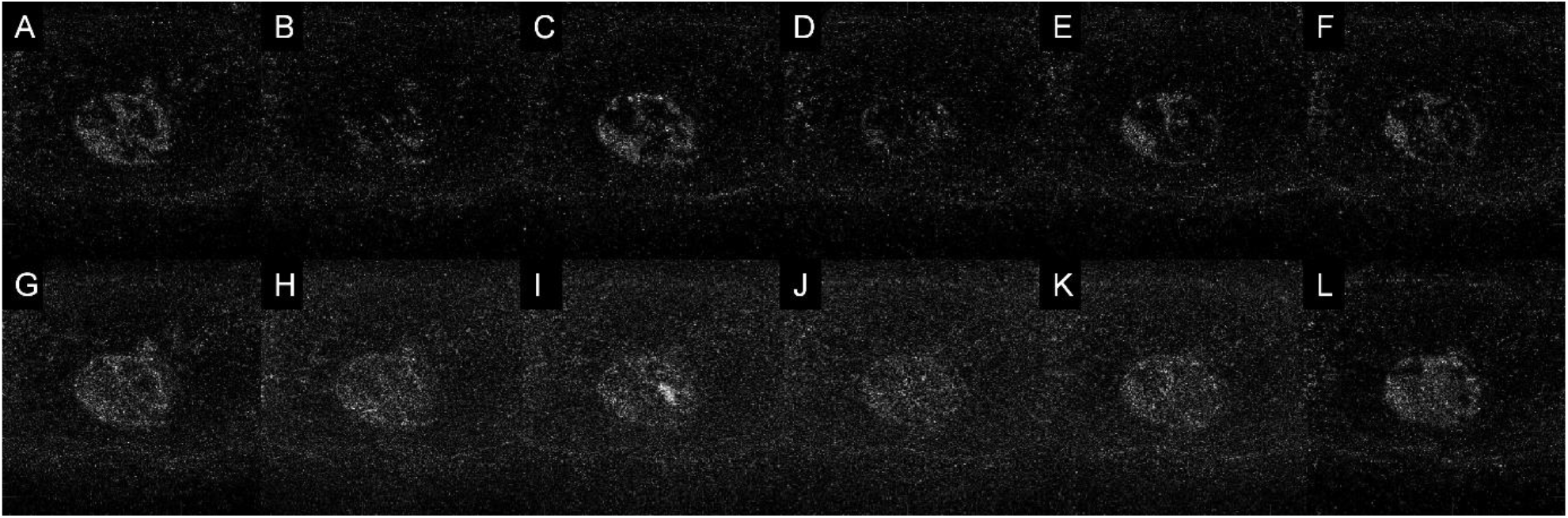
All subfigures are in transverse plane and display some registration residuals (difference between a moving and a reference image) of registrations performed on the same XD-GRASP_4D+ reconstruction. The top row displays the residuals of registrations performed between a frame and its previous neighbor like in MASTER_3D+. The bottom row displays the residuals of registrations performed between the same frame as in the top row but with the first frame as reference image (“all-to-one” registration). Visual inspection shows that the registration residuals between neighboring frames is sparser than the registration residuals between the same frames and the first frame.

## References

1. Di--Sopra L, Piccini D, Coppo S, Stuber M, Yerly J. An automated approach to fully self-gated free-running cardiac and respiratory motion-resolved 5D whole-heart MRI. Magn Reson Med. 2019 Dec;82(6):2118–32.

2. Van--Den--Hout, RJ, Lamb HJ, Van--Den--Aardweg, JG, Schot R, Steendijk P, Van--Der--Wall, EE, et al. Real-Time MR Imaging of Aortic Flow: Influence of Breathing on Left Ventricular Stroke Volume in Chronic Obstructive Pulmonary Disease. Radiology. 2003 Nov;229(2):513–9.

3. Usman M, Ruijsink B, Nazir MS, Cruz G, Prieto C. Free breathing whole-heart 3D CINE MRI with self-gated Cartesian trajectory. Magn Reson Imaging. 2017 May;38:129–37.

4. Küstner T, Bustin A, Jaubert O, Hajhosseiny R, Masci PG, Neji R, et al. Fully self-gated free-running 3D Cartesian cardiac CINE with isotropic whole-heart coverage in less than 2 min. NMR Biomed. 2021 Jan;34(1):e4409.

5. Kozerke S, Tsao J, Razavi R, Boesiger P. Accelerating cardiac cine 3D imaging using k-t BLAST. Magn Reson Med. 2004 Jul;52(1):19–26.

6. Jung H, Ye JC, Kim EY. Improved k-t BLAST and k-t SENSE using FOCUSS. Phys Med Biol. 2007 Jun 7;52(11):3201–26.

7. Tsao J, Boesiger P, Pruessmann KP. k-t BLAST and k-t SENSE: dynamic MRI with high frame rate exploiting spatiotemporal correlations. Magn Reson Med. 2003 Nov;50(5):1031–42.

8. Hansen MS, Kozerke S, Pruessmann KP, Boesiger P, Pedersen EM, Tsao J. On the influence of training data quality in k-t BLAST reconstruction. Magn Reson Med. 2004 Nov;52(5):1175–83.

9. Tsao J, Kozerke S, Boesiger P, Pruessmann KP. Optimizing spatiotemporal sampling for k-t BLAST and k-t SENSE: application to high-resolution real-time cardiac steady-state free precession. Magn Reson Med. 2005 Jun;53(6):1372–82.

10. Gamper U, Boesiger P, Kozerke S. Compressed sensing in dynamic MRI. Magn Reson Med. 2008 Feb;59(2):365–73.

11. Usman M, Prieto C, Odille F, Atkinson D, Schaeffter T, Batchelor PG. A computationally efficient OMP-based compressed sensing reconstruction for dynamic MRI. Phys Med Biol. 2011 Apr 7;56(7):N99–114.

12. Royuela-del-Val J, Cordero-Grande L, Simmross-Wattenberg F, Martín-Fernández M, Alberola-López C. Jacobian weighted temporal total variation for motion compensated compressed sensing reconstruction of dynamic MRI: Jacobian Weighted temporal Total Variation. Magn Reson Med. 2017 Mar;77(3):1208–15.

13. Asif MS, Hamilton L, Brummer M, Romberg J. Motion-adaptive spatio-temporal regularization for accelerated dynamic MRI. Magn Reson Med. 2013 Sep;70(3):800–12.

14. Liu F, Li D, Jin X, Qiu W, Xia Q, Sun B. Dynamic cardiac MRI reconstruction using motion aligned locally low rank tensor (MALLRT). Magn Reson Imaging. 2020 Feb;66:104–15.

15. Menchón-Lara, RM, Royuela-del-Val J, Godino-Moya A, Cordero-Grande L, Simmross-Wattenberg F, Martín-Fernández M, et al. An Efficient Multi-resolution Reconstruction Scheme with Motion Compensation for 5D Free-Breathing Whole-Heart MRI. In: Cardoso MJ, Arbel T, Gao F, Kainz B, van Walsum, T, Shi K, et al., editors. Molecular Imaging, Reconstruction and Analysis of Moving Body Organs, and Stroke Imaging and Treatment [Internet]. Cham: Springer International Publishing; 2017 [cited 2022 Oct 22]. p. 136–45. (Lecture Notes in Computer Science; vol. 10555). Available from: http://link.springer.com/10.1007/978-3-319-67564-0_14

16. Pang J, Chen Y, Fan Z, Nguyen C, Yang Q, Xie Y, et al. High efficiency coronary MR angiography with nonrigid cardiac motion correction. Magn Reson Med. 2016 Nov;76(5):1345–53.

17. Mohsin YQ, Lingala SG, DiBella E, Jacob M. Accelerated dynamic MRI using patch regularization for implicit motion compensation. Magn Reson Med. 2017 Mar;77(3):1238–48.

18. Phair A, Cruz G, Qi H, Botnar RM, Prieto C. Free-running 3D whole-heart T1 and T2 mapping and cine MRI using low-rank reconstruction with non-rigid cardiac motion correction. Magn Reson Med. 2023 Jan;89(1):217–32.

19. Godino-Moya A, Royuela-del-Val J, Usman M, Menchón-Lara, RM, Martín-Fernández M, Prieto C, et al. Space-time variant weighted regularization in compressed sensing cardiac cine MRI. Magnetic Resonance Imaging. 2019 May;58:44–55.

20. Tolouee A, Alirezaie J, Babyn P. Nonrigid motion compensation in compressed sensing reconstruction of cardiac cine MRI. Magn Reson Imaging. 2018 Feb;46:114–20.

21. Jung H, Sung K, Nayak KS, Kim EY, Ye JC. k-t FOCUSS: A general compressed sensing framework for high resolution dynamic MRI: k-t FOCUSS. Magn Reson Med. 2009 Jan;61(1):103–16.

22. Prieto C, Batchelor PG, Hill DLG, Hajnal JV, Guarini M, Irarrazaval P. Reconstruction of undersampled dynamic images by modeling the motion of object elements. Magn Reson Med. 2007 May;57(5):939–49.

23. Jung H, Ye JC. Motion estimated and compensated compressed sensing dynamic magnetic resonance imaging: What we can learn from video compression techniques. Int J Imaging Syst Technol. 2010 May 19;20(2):81–98.

24. Chen X, Salerno M, Yang Y, Epstein FH. Motion-compensated compressed sensing for dynamic contrast-enhanced MRI using regional spatiotemporal sparsity and region tracking: block low-rank sparsity with motion-guidance (BLOSM). Magn Reson Med. 2014 Oct;72(4):1028–38.

25. Usman M, Atkinson D, Odille F, Kolbitsch C, Vaillant G, Schaeffter T, et al. Motion corrected compressed sensing for free-breathing dynamic cardiac MRI. Magn Reson Med. 2013 Aug;70(2):504–16.

26. Bustin A, Rashid I, Cruz G, Hajhosseiny R, Correia T, Neji R, et al. 3D whole-heart isotropic sub-millimeter resolution coronary magnetic resonance angiography with non-rigid motion-compensated PROST. J Cardiovasc Magn Reson. 2020 Dec;22(1):24.

27. Zhao N, O’Connor D, Basarab A, Ruan D, Sheng K. Motion Compensated Dynamic MRI Reconstruction With Local Affine Optical Flow Estimation. IEEE Trans Biomed Eng. 2019 Nov;66(11):3050–9.

28. Lingala SG, DiBella E, Jacob M. Deformation Corrected Compressed Sensing (DC-CS): A Novel Framework for Accelerated Dynamic MRI. IEEE Trans Med Imaging. 2015 Jan;34(1):72–85.

29. Correia T, Ginami G, Cruz G, Neji R, Rashid I, Botnar RM, et al. Optimized respiratory-resolved motion-compensated 3D Cartesian coronary MR angiography. Magn Reson Med. 2018 Dec;80(6):2618–29.

30. Royuela-del-Val J, Cordero-Grande L, Simmross-Wattenberg F, Martín-Fernández M, Alberola-López C. Nonrigid groupwise registration for motion estimation and compensation in compressed sensing reconstruction of breath-hold cardiac cine MRI: Groupwise ME/MC in CS Reconstruction of Cardiac Cine MRI. Magn Reson Med. 2016 Apr;75(4):1525–36.

31. Coppo S, Piccini D, Bonanno G, Chaptinel J, Vincenti G, Feliciano H, et al. Free-running 4D whole-heart self-navigated golden angle MRI: Initial results. Magn Reson Med. 2015 Nov;74(5):1306–16.

32. Vincenti G, Monney P, Chaptinel J, Rutz T, Coppo S, Zenge MO, et al. Compressed Sensing Single–Breath-Hold CMR for Fast Quantification of LV Function, Volumes, and Mass. JACC: Cardiovascular Imaging. 2014 Sep;7(9):882–92.

33. Feng L, Axel L, Chandarana H, Block KT, Sodickson DK, Otazo R. XD-GRASP: Golden-angle radial MRI with reconstruction of extra motion-state dimensions using compressed sensing. Magn Reson Med. 2016 Feb;75(2):775–88.

34. Lustig M, Donoho D, Pauly JM. Sparse MRI: The application of compressed sensing for rapid MR imaging. Magn Reson Med. 2007 Dec;58(6):1182–95.

35. Hestenes MR, Stiefel E. Methods of conjugate gradients for solving linear systems. J RES NATL BUR STAN. 1952 Dec;49(6):409.

36. Li C, Yin W, Jiang H, Zhang Y. An efficient augmented Lagrangian method with applications to total variation minimization. Comput Optim Appl. 2013 Dec;56(3):507–30.

37. Qiao Z, Redler G, Epel B, Halpern H. A Simple but Universal Fully Linearized ADMM Algorithm for Optimization Based Image Reconstruction [Internet]. In Review; 2023 Apr [cited 2023 Aug 6]. Available from: https://www.researchsquare.com/article/rs-2857384/v1

38. Roy CW, Di Sopra L, Whitehead KK, Piccini D, Yerly J, Heerfordt J, et al. Free-running cardiac and respiratory motion-resolved 5D whole-heart coronary cardiovascular magnetic resonance angiography in pediatric cardiac patients using ferumoxytol. J Cardiovasc Magn Reson. 2022 Dec;24(1):39.

39. Masala N, Bastiaansen JAM, Di Sopra L, Roy CW, Piccini D, Yerly J, et al. Free-running 5D coronary MR angiography at 1.5T using LIBRE water excitation pulses. Magn Reson Med. 2020 Sep;84(3):1470–85.

40. Milani B. A mathematical Language for MRI Reconstructions’, version 0.0 [Internet]. 2023. Available from: https://drive.google.com/file/d/12z9JCFhwBJhDW4_3Uy4bhSXCnvPod0os/view?usp=sharing

41. Pruessmann KP, Weiger M, Börnert P, Boesiger P. Advances in sensitivity encoding with arbitrary k-space trajectories: SENSE With Arbitrary k-Space Trajectories. Magn Reson Med. 2001 Oct;46(4):638–51.

42. Piccini D, Littmann A, Nielles-Vallespin S, Zenge MO. Spiral phyllotaxis: The natural way to construct a 3D radial trajectory in MRI: Spiral Phyllotaxis Radial 3D Trajectory. Magn Reson Med. 2011 Oct;66(4):1049–56.

43. Boyd S. Distributed Optimization and Statistical Learning via the Alternating Direction Method of Multipliers. FNT in Machine Learning. 2010;3(1):1–122.

44. Marc Modat. NiftyReg [Internet]. Available from: https://sourceforge.net/projects/niftyreg/

45. Modat M, Ridgway GR, Taylor ZA, Lehmann M, Barnes J, Hawkes DJ, et al. Fast free-form deformation using graphics processing units. Computer Methods and Programs in Biomedicine. 2010 Jun;98(3):278–84.

46. Etienne A, Botnar RM, van Muiswinkel, AMC, Boesiger P, Manning WJ, Stuber M. ‘Soap-Bubble’ visualization and quantitative analysis of 3D coronary magnetic resonance angiograms. Magn Reson Med. 2002 Oct;48(4):658–66.

47. Ahmad R, Ding Y, Simonetti OP. Edge sharpness assessment by parametric modeling: Application to magnetic resonance imaging: EDGE SHARPNESS ASSESSMENT FOR MRI. Concepts Magn Reson. 2015 May;44(3):138–49.

48. Laubrock K, Von Loesch T, Steinmetz M, Lotz J, Frahm J, Uecker M, et al. Imaging of arrhythmia: Real-time cardiac magnetic resonance imaging in atrial fibrillation. European Journal of Radiology Open. 2022;9:100404.

49. Roy CW, Heerfordt J, Piccini D, Rossi G, Pavon AG, Schwitter J, et al. Motion compensated whole-heart coronary cardiovascular magnetic resonance angiography using focused navigation (fNAV). J Cardiovasc Magn Reson. 2021 Dec;23(1):33.

50. Segars WP, Sturgeon G, Mendonca S, Grimes J, Tsui BMW. 4D XCAT phantom for multimodality imaging research. Med Phys. 2010 Sep;37(9):4902–15.

51. Wissmann L, Santelli C, Segars WP, Kozerke S. MRXCAT: Realistic numerical phantoms for cardiovascular magnetic resonance. J Cardiovasc Magn Reson. 2014 Dec;16(1):63.

52. Lingala SG, Hu Y, DiBella E, Jacob M. Accelerated Dynamic MRI Exploiting Sparsity and Low-Rank Structure: k-t SLR. IEEE Trans Med Imaging. 2011 May;30(5):1042–54.

53. Pruessmann KP, Weiger M, Börnert P, Boesiger P. Advances in sensitivity encoding with arbitrary k-space trajectories: SENSE With Arbitrary k-Space Trajectories. Magn Reson Med. 2001 Oct;46(4):638–51.

54. Milani B. A mathematical Language for MRI Reconstructions’, version 0.0 [Internet]. 2023. Available from: https://drive.google.com/file/d/12z9JCFhwBJhDW4_3Uy4bhSXCnvPod0os/view?usp=sharing

55. Walsh DO, Gmitro AF, Marcellin MW. Adaptive reconstruction of phased array MR imagery. Magn Reson Med. 2000 May;43(5):682–90.

56. Griswold M, Walsh D, Heidemann R, Haase A, Jakob P. The Use of an Adaptive Reconstruction for Array Coil Sensitivity Mapping and Intensity Normalization. Proc. Intl. Soc. Mag. Reson. Med.; 2002.

57. Herterich R, Sumarokova A. Coil Sensitivity Estimation and Intensity Normalisation for Magnetic Resonance Imaging. 2019 [cited 2023 Apr 15]; Available from: 10.13140/RG.2.2.22886.04169

58. Royuela-del-Val J, Cordero-Grande L, Simmross-Wattenberg F, Martín-Fernández M, Alberola-López C. Nonrigid groupwise registration for motion estimation and compensation in compressed sensing reconstruction of breath-hold cardiac cine MRI: Groupwise ME/MC in CS Reconstruction of Cardiac Cine MRI. Magn Reson Med. 2016 Apr;75(4):1525–36.

59. Menchón-Lara, RM, Royuela-del-Val J, Godino-Moya A, Cordero-Grande L, Simmross-Wattenberg F, Martín-Fernández M, et al. An Efficient Multi-resolution Reconstruction Scheme with Motion Compensation for 5D Free-Breathing Whole-Heart MRI. In: Cardoso MJ, Arbel T, Gao F, Kainz B, van Walsum, T, Shi K, et al., editors. Molecular Imaging, Reconstruction and Analysis of Moving Body Organs, and Stroke Imaging and Treatment [Internet]. Cham: Springer International Publishing; 2017 [cited 2022 Oct 22]. p. 136–45. (Lecture Notes in Computer Science; vol. 10555). Available from: http://link.springer.com/10.1007/978-3-319-67564-0_14

